# Neuropsychology of chronic back pain managed with long-term opioid use

**DOI:** 10.1101/2024.02.07.24302408

**Authors:** Marwan N Baliki, Andrew D. Vigotsky, Gaelle Rached, Rami Jabakhanji, Lejian Huang, Paulo Branco, Olivia Cong, James Griffith, Ajay D. Wasan, Thomas J. Schnitzer, A. Vania Apkarian

## Abstract

Chronic pain is commonly treated with long-term opioids, but the neuropsychological outcomes associated with stable long-duration opioid use remain unclear. Here, we contrasted the psychological profiles, brain activity, and brain structure of 70 chronic back pain patients on opioids (CBP+O, average opioid exposure 6.2 years) with 70 patients managing their pain without opioids. CBP+O exhibited moderately worse psychological profiles and small differences in brain morphology. However, CBP+O had starkly different spontaneous brain activity, dominated by increased mesocorticolimbic and decreased dorsolateral-prefrontal activity, even after controlling for pain intensity and duration. These differences strongly reflected cortical opioid and serotonin receptor densities and mapped to two antagonistic resting-state circuits. The circuits’ dynamics were explained by mesocorticolimbic activity and reflected negative affect. We reassessed a sub-group of CBP+O after they briefly abstained from taking opioids. Network dynamics, but not spontaneous activity, reflected exacerbated signs of withdrawal. Our results have implications for the management and tapering of opioids in chronic pain.

## Main

Chronic pain afflicts over 20% of the world’s population ^1^, and has massive economic and societal ramifications. Debilitating pain can hijack patients’ lives, preventing them from freely being able to work, exercise, and socialize. Chronic low back pain (CBP), for example, is the leading cause of disability in the world ^2^. Until the mid-2010s, prescription opioids were often used as a first-line treatment for chronic pain, making chronic pain a natural entry point for opioid use and, thus, potential downstream misuse, opioid use disorder (OUD), and even overdose ^3–5^. Despite marked decreases in opioid prescription rates for chronic pain over the past ten years, opioids are still used by 20–50% of patients suffering from chronic pain ^6–8^. The neuropsychological implications of long-term opioid use in patients with chronic pain are largely unknown.

Chronic pain and opioids have been extensively studied on their own. Each imparts a massive societal cost ^9,10^, and their combination may impart an even greater cost. Our group has demonstrated that the development of chronic pain involves mesocorticolimbic plasticity, including the limbic amygdala, hippocampus, nucleus accumbens (NAc), and associated cortical circuits: medial prefrontal cortex (mPFC) and posterior cingulate, precuneus cortex ^11–16^. Interestingly, these same circuits are strongly implicated in OUD, especially the dopaminergic ventral tegmental area (VTA), NAc, and mPFC circuit ^17,18^. From a high level, mesocorticolimbic circuits help code emotional valence and modulate behavioral responses, such as appetition and aversion ^19–21^. Pain is an aversive experience—patients actively avoid triggers. In contrast, opioids, while commonly said to be rewarding, may induce aversion and negative affect during periods of withdrawal—so-called hyperkatifeia ^22^. As a result, the mechanisms of chronic pain may interact with long-term opioid use, exacerbating patients’ pain experience when off opioids to create a vicious cycle. However, there is little data on the impact of long-term opioid consumption in chronic pain patients ^23–28^.

In this work, we aimed to answer three questions: **(1)** Is long-term opioid prescription use in patients with chronic pain associated with better (or worse) clinical outcomes? **(2)** Is long-term opioid use in patients with chronic pain associated with differential brain activity and structure, especially across the mesocorticolimbic system? And **(3)** how do the results of **(1)** and **(2)** interrelate? To answer these questions, we compared the clinical, psychological, and brain anatomical and functional properties of CBP on long-term stable doses of opioids (CBP+O, n=70; mean = 6.2 years of opioid consumption) and patients managing their pain without opioids (CBP−O, n=70). The two groups were one-to-one matched (from a 150 CBP−O dataset) for age, sex, pain intensity (mean = 5.4 on a 0–10 numeric rating scale), and pain duration (>0.5 years, mean = 17 years) (**Extended Data Table 1**). We reasoned that contrasting the neuropsychological characteristics of CBP+O and CBP−O – two patient groups with closely matched demographics and pain characteristics except for opioid use – would allow us to begin to clinically and mechanistically understand long-term opioid consumption in patients with chronic pain.

## Results

### Clinical and psychological characteristics of chronic pain patients with long-term opioid use

Chronic pain and opioid use are independently associated with multiple psychological comorbidities, including negative affect, sleep disturbance, and diminished social interactions ^5,18,29^. Are these outcomes worse in patients managing their chronic pain with long-term opioid consumption? Currently, there is little data to answer this question ^23,30,31^. There is clinical equipoise: On one hand, prescription opioids may improve patients’ mental health via pain relief. On the other hand, if prescription opioids enhance the likelihood of OUD, they may exacerbate rather than relieve the psychological challenges associated with chronic pain, for example, prescription opioids are associated with worse depression ^32^.

We assessed the clinical and psychological characteristics of CBP patients, for which we used validated self-report questionnaires (**Fig. 1a**). This allowed us to compare how patients on long-term opioid therapy fare clinically and psychologically relative to patients without opioids. To this end, we assessed patients’ physical function, pain, and mental health using 17 measures. We performed a principal components analysis (PCA) to reduce the dimensionality of these outcomes. Three principal components (PC1–3; > 2, variance > 10%) explained 69% of the total variance: PC1 (which we call *functional disability*) included decreased physical function, less social activity, increased general disability, greater pain interference, and greater fatigue; PC2 (*pain quality*) was mainly composed of pain properties, including sensory and affective pain scales, neuropathic pain symptoms, and pain catastrophizing; PC3 (*negative affect*) included increased depression, anxiety, negative affect, decreased mental well-being, and less sleep (**Fig. 1a, Extended Data Table 2**). These PCs were stable no matter how they were derived (**Extended Data Fig. 1, Extended Data Table 2)** and closely resemble the results we have previously reported ^23^. These PCs were used to infer group (CBP+O vs. CBP−O) differences in clinical, psychological, and brain characteristics.

**Fig. 1.**
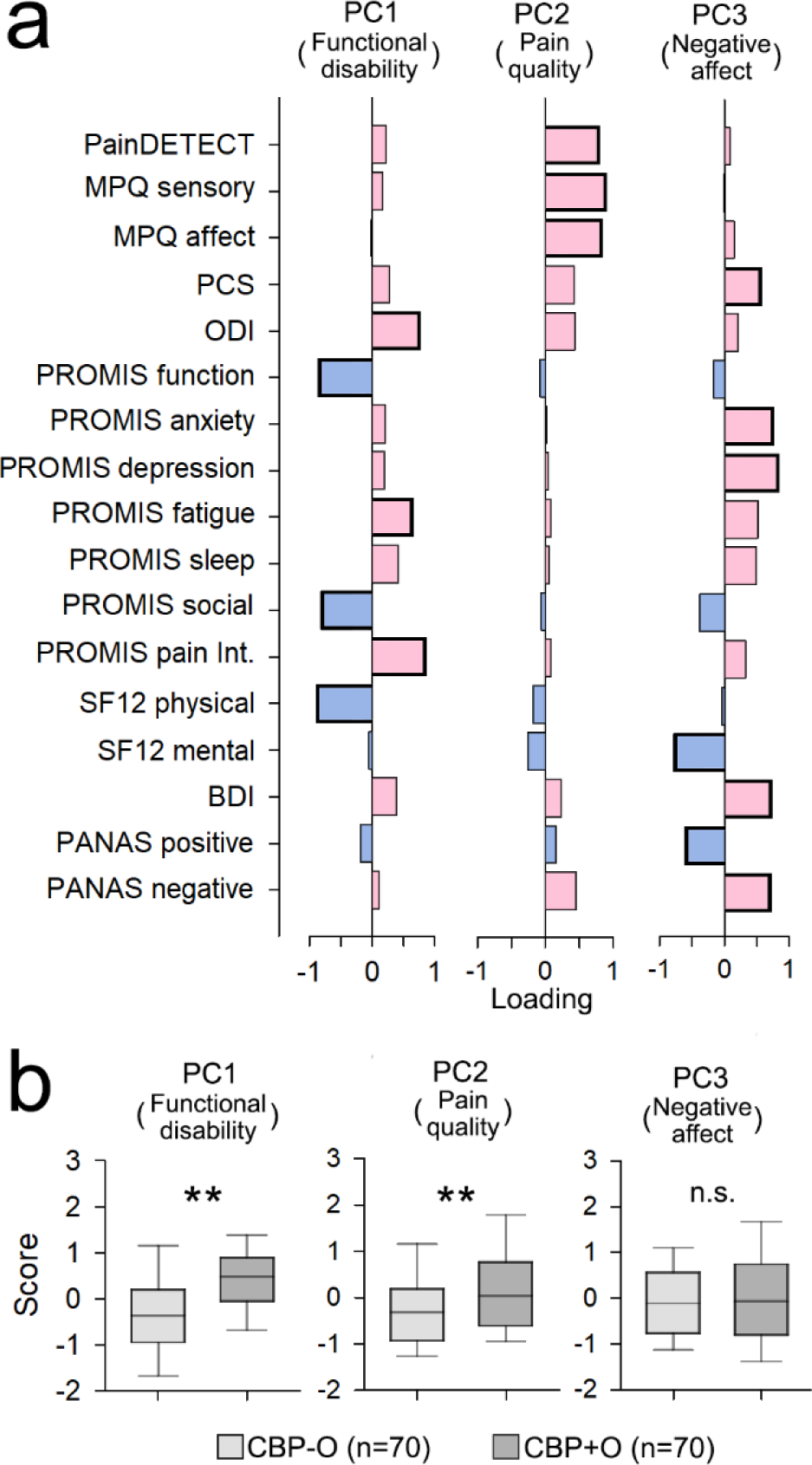
Long-term opioid use was associated with worse functional disabilities and worse pain quality. **a.** Clinical parameters loaded on three main components that explained 69.9% of the variance, using principal components analysis (PCA). Bars represent factor loadings of each measure on each principal component (PC). Identified PC-s were uniquely associated with functional disability (PC1), pain quality (PC2), and negative affect (PC3). Blue bars represent measurements where lower scores correspond to worse outcomes. Red bars represent measurements where higher scores correspond to worse outcomes. Bolded outlines represent large loading factors (>0.5). **b.** Compared to CBP-O, CBP+O exhibited higher functional disability (PC1, p<0.01) and worse pain quality (PC2, p<0.01). PainDETECT = presence of neuropathic pain; MPQ = McGill Pain Questionnaire, subscales are indicated; PCS = Pain Catastrophizing Scale; ODI = Oswestry low back Disability Index; PROMIS = Patient-Reported Outcomes Measurement Information System, subscales are labeled, pain int. = pain interference; SF12 = Short Form quality of life scale; BDI = Becks Depression Inventory; PANAS = Positive and Negative Affect Schedule.

Group (CBP+O vs. CBP−O) differences for the three PC scores were assessed using analysis of covariance (ANCOVA) with sex, race, age, pain intensity (NRS), pain duration (log), body mass index (BMI), and medication quantification scale (MQS) included as covariates. CBP+O exhibited higher functional disability (PC1) and worse pain quality (PC2) scores compared to CBP−O. The groups had similar negative affect (PC3), but levels of depression (BDI scores) were twice as high in CBP+O relative to CBP−O (**Fig. 1b, Extended Data Table 3**). The 17 measures comprising the PCs largely mirrored the PC results—CBP+O had statistically significantly poorer outcomes for 7 of the 17 measures; only anxiety (PROMIS) was similar, and all other outcomes tended to show worse values in CBP+O relative to CBP−O, albeit with appreciable uncertainty (**Extended Data Table 4**). Although negative affect (PC3) did not statistically significantly differ between the groups, we have previously shown that CBP−O exhibits poorer outcomes in comparison to CBP+O on all five emotional scales of the NIH Toolbox assessment, especially on the negative affect scale ^31^.

Even though back pain intensity was matched between CBP−O and CBP+O, and PC2 was correlated with back pain in both groups, the consumption of non-opioid medications (MQS) was twice that in CBP+O compared to CBP−O and correlated with back pain intensity only in CBP+O (**Extended Data Fig. 2**). Overall, the clinical phenotyping indicates that patients on stable, long-term prescription opioid treatment 1) exhibit worse pain qualities, particularly neuropathic-like pain with a larger sensory component; 2) show higher functional disability; 3) do not differ in negative affect from non-opioid users; and 4) consume more non-opioid medications than those not on opioids, in proportion with their reported pain intensity. Although opioid consumption was not associated with an improvement in any of the 17 clinical parameters examined, it is essential to note that many of the differences between CBP+O and CBP−O were modest.

### Relationships between opioid use outcomes and the clinical and psychological characteristics of patients with chronic back pain with long-term opioid use

Opioid dosage is a strong determinant of patient safety and is considered when deciding to taper^33^. We examined how opioid use in CBP+O relates to clinical measures, including withdrawal symptoms and misuse risk. Opioid use was quantified using three measurements: 1) the daily prescription converted into morphine milligram equivalent (MME); 2) blood levels of opioids converted to a relative opioid equivalent (ROE, mg/L); and 3) the duration of opioid use (DOU) (**Fig. 2a**). MME was used to subdivide CBP+O by OUD risk as defined by the Centers for Disease Control and Prevention (CDC) guidelines ^34^ (**Fig. 2a** top panel). All three measures—MME, ROE, and DOU—were right-skewed and thus log-transformed. Blood opioids (ROE) in CBP+O reflected their prescription dose (MME) (*r*=0.48, *p*<0.001; **Fig. 2b** scatter plot), but neither blood opioid levels (ROE) nor prescription dose (MME) was strongly associated with the duration of opioid use or with non-opioid medication use (MQS) (**Fig. 2b**).

**Fig. 2.**
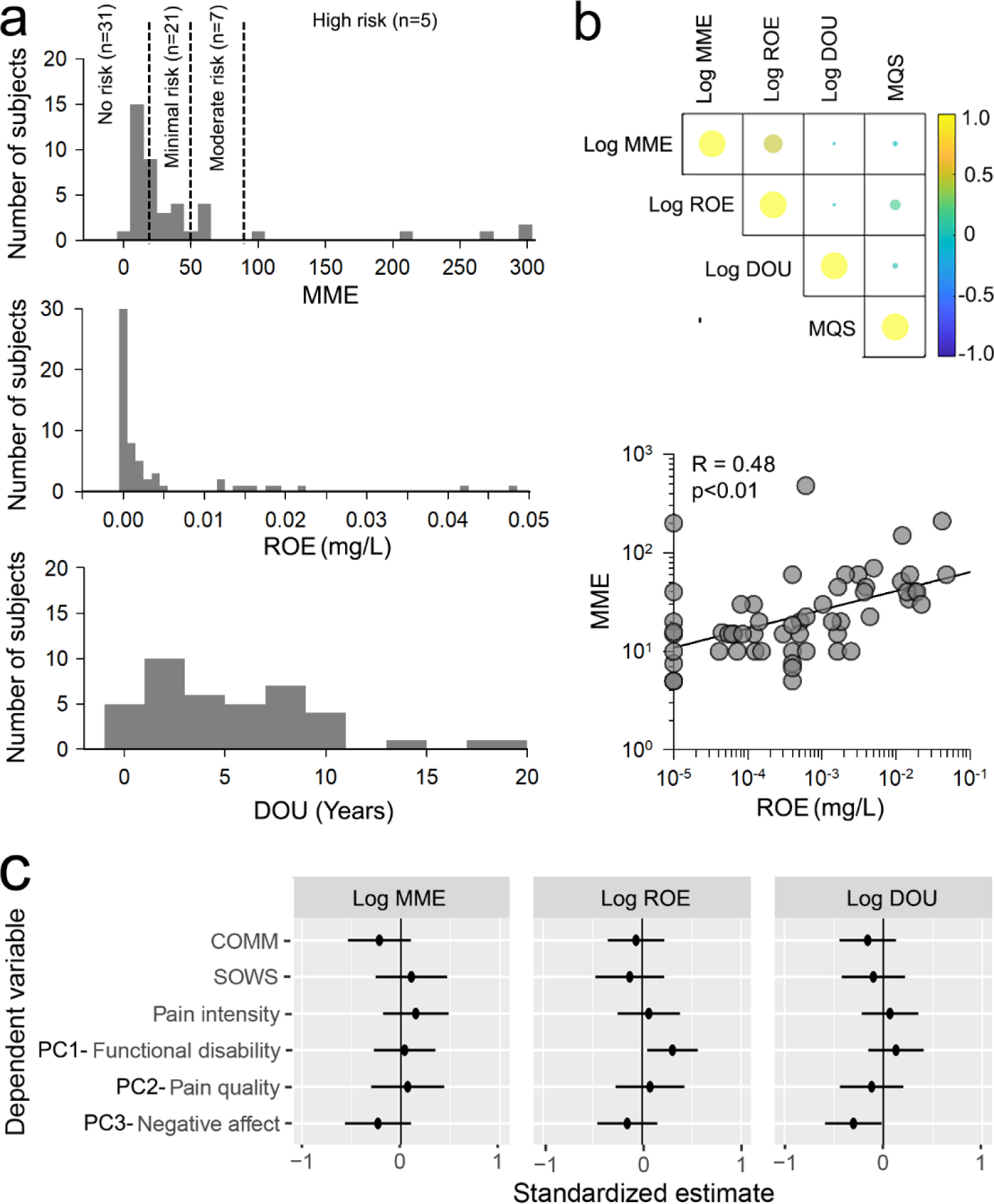
Opioid measurements and their relationship to clinical and psychological parameters in CBP patients on long-term opioids (CBP+O). **a.** Distribution of opioid prescription consumption (MME), blood opioid levels (ROE in mg/L), and duration of opioid use (DOU in years) in CBP+O. CBP+O showed a varied and wide range of opioid use, with 31 patients within the no-risk range (MME < 20), 21 patients within the minimal risk range (MME 20-50), 7 patients within the moderate risk range (MME 50-90) and 5 patients within the high-risk range (MME > 90). All three outcomes (MME, ROE, DOU) were right-skewed. Therefore, they were log-transformed to conform to normal distributions. **b.** The heat map shows the relationship of opioid measures with each other, and with consumption of non-opioid medications (MQS). Only MME and ROE showed a significant relationship (scatter plot). **c.** The plot shows standardized regression estimates (± 95% CI) for the relationship between opioid measures (log) with signs of misuse (COMM) and withdrawal (SOWS) scales, pain intensity, and the three principal components that characterize the population across all CBP+O. Only opioid blood levels (ROE) showed a significant relationship with PC1-functional disability.

We assessed patients’ withdrawal symptoms and misuse risk using validated scales (subjective opiate withdrawal scale, SOWS; current opioid misuse measure, COMM). On these scales, 19 CBP+O (27%) showed high misuse risk (COMM > 9) and 7 (10%) high withdrawal symptoms (SOWS > 20). We investigated these scales’ relationships with back pain intensity, functional disability (PC1), pain quality (PC2), and negative affect (PC3). Opioid use characteristics (MME, ROE, and DOU) were not consistently associated with withdrawal or misuse. Only ROE showed a statistically significant positive correlation with functional disability (*r*=0.47, *p*<0.01; **Extended Data Fig. 3**). There was no statistically significant association between opioid usage parameters with pain intensity, pain quality, or negative affect scores (PC1–3, **Fig. 2c**). Finally, subdividing CBP+O patients into low (MME < 50) and high (MME ≥ 50) opioid consumers did not yield remarkable differences in the correlations between clinical and opioid parameters (**Extended Data Fig. 4**). Thus, CBP+O who consumed more opioids were not necessarily at greater risk of OUD.

### Brain morphology of chronic pain patients with long-term opioid use

Chronic pain and OUD are each associated with global and local reorganization in brain gray matter properties ^35–38^. We computed normalized peripheral gray matter volumes (PGMV) for all participants (see *Methods*). After adjusting for covariates of no interest, CBP+O showed lower PGMV than CBP−O (*p*<0.05) (**Fig. 3a, Extended Data Table 5**). However, PGMV was not related to opioid use (MME, ROE, and DOU), clinical parameters (PC1–3), or withdrawal and misuse scales (**Extended Data Table 6).** We also investigated subcortical volumes (see *Methods*), which did not statistically significantly differ between CBP+O and CBP−O (**Extended Data Fig 6, Extended Data Table 7).**

**Fig. 3.**
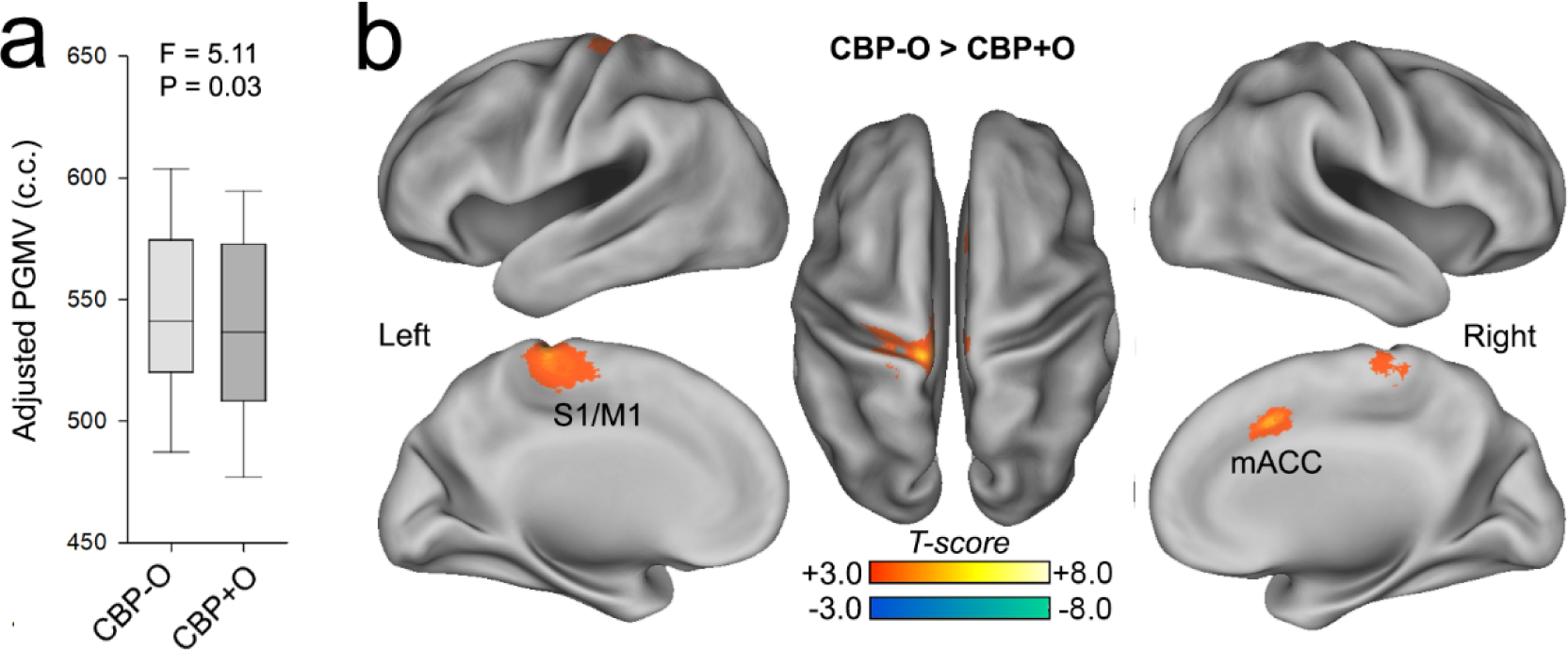
Long-term opioid use was associated with lower global and local gray matter. **a.** Peripheral gray matter volume (PGMV) was significantly less in CBP+O compared to CBP-O. **b.** CBP+O showed less gray matter density in S1/M1 and mACC (TFCC t-score > 2.3, p<0.01 corrected for multiple comparisons). PGMV = peripheral gray matter volume; S1/M1 = primary sensorimotor cortex; mACC = middle anterior cingulate cortex.

We used whole-brain voxel-based morphometry (VBM) to investigate regional grey matter density (GMD) differences between CBP+O and CBP−O. After adjusting for covariates of no interest, we found two clusters localized to (1) the left primary sensorimotor cortex (S1/M1) and (2) the mid-anterior cingulate cortex (mACC), which showed decreased GMD in CBP+O. We used reverse inference in Neurosynth to identify the top 5 terms (out of 1,765) associated with each cluster. The S1/M1 cluster was associated with sensorimotor function, while mACC with pain, nociception, and arousal (**Fig. 3b, Extended Data Table 8**). In addition, mACC GMD was negatively correlated with both back pain intensity (*p*=0.02) and duration (*p*=0.01) in both groups (**Extended data Fig. 7, Extended Data Table 9**). Similar to our PGMV analysis, localized GMD changes in mACC and S1/M1 were not statistically significantly associated with opioid use, withdrawal, misuse, or clinical parameters (PC1–3) in CBP+O patients (**Extended data table 10**). Overall, whole-brain gray matter decreases were modest, and focal decreases in cortical gray matter volume were associated with long-term opioid use in CBP+O.

To examine the impact of opioid exposure on the white matter, we contrasted the 58 CBP+O and matched 58 CBP−O’s white matter properties using a whole-brain skeletal fractional anisotropy (FA) contrast, which did not yield any statistically significant differences between the two groups (data not shown).

### Changes in spontaneous brain activity in chronic pain patients with long-term opioid use

The power spectrum of brain activity signals is related to various brain functional properties ^39,40^. We used resting-state fMRI to localize whole-brain voxel-wise differences in the amplitude of low-frequency fluctuations (ALFF), which reflects spontaneous neural activity ^41^, between CBP+O and CBP−O while adjusting for age, sex, pain intensity, pain duration, BMI, MQS, and head motion. We also included voxel-wise corrections for scanner signal-to-noise ratios and GMD values. Compared to CBP−O, CBP+O showed increased ALFF in five distinct clusters, the largest of which (5,050 voxels) encompassed multiple mesocorticolimbic structures, including bilateral nucleus accumbens (NAc), amygdala, subgenual cingulate, orbital prefrontal cortex, hippocampus, and brain stem. Other clusters that showed increased ALFF in CBP+O included the lateral occipital cortex, the middle prefrontal cortex, and the right and left posterior portions of the inferior temporal gyrus. CBP+O patients also showed decreased ALFF in 3 clusters, including the left dorsolateral prefrontal cortex (dlPFC, 3,955 voxels) and the right and left anterior part of the mid-temporal gyrus (**Fig 4a, Extended Table 11**). Increased ALFF in CBP+O was localized to brain regions involved in reward, motivation, incentive, and value processing, while decreased ALFF in CBP+O was localized to brain regions involved in language and mental states **(Fig 4b).**

**Fig. 4.**
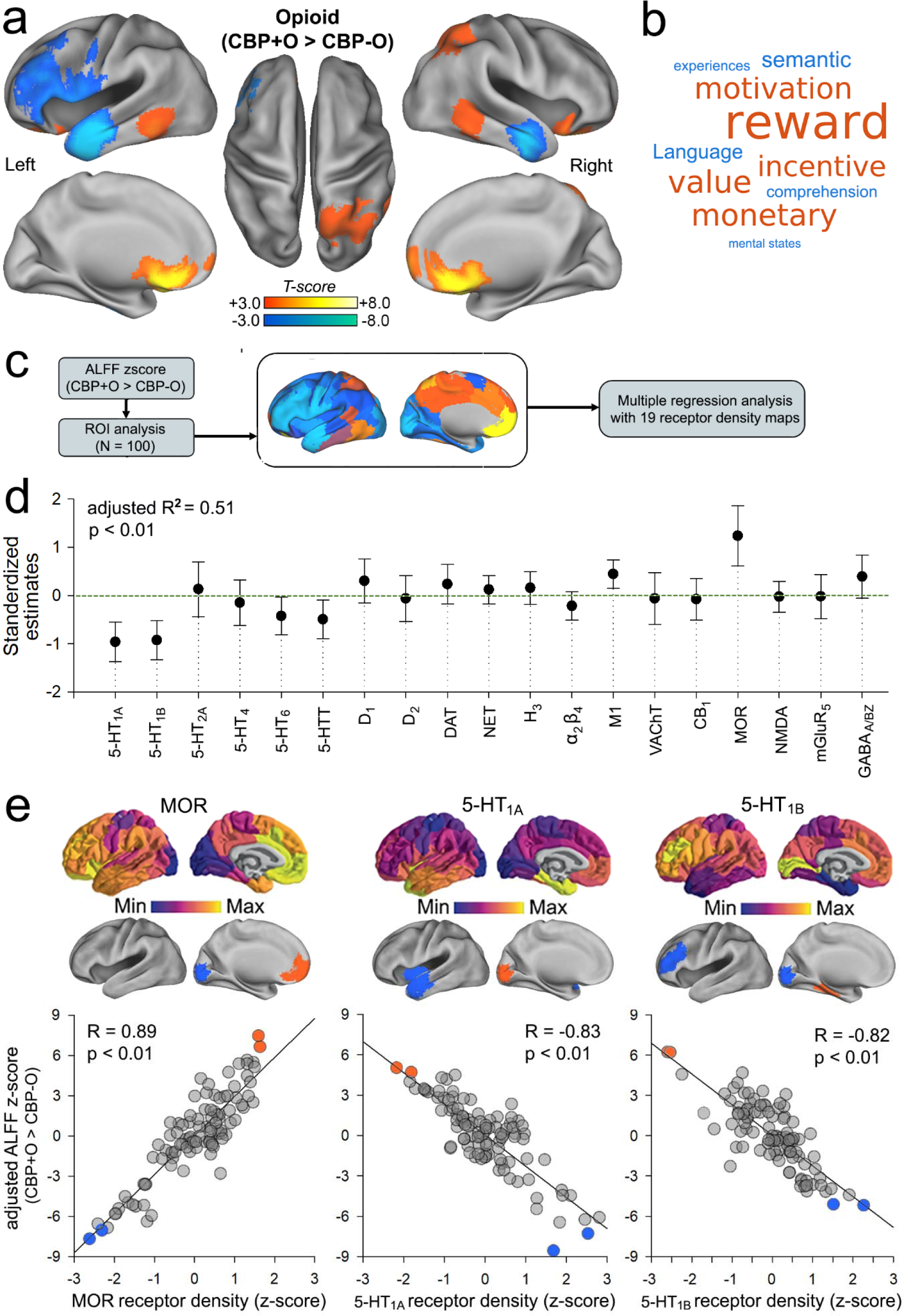
Spontaneous activity with long-term opioid use mapped to specific cortical receptor density distributions. **a.** Brain images show regions that showed a significant increase (red-yellow) and decreased (blue-green) spontaneous activity (change in ALFF) in CBP+O compared to CBP-O. Regions that showed increased activity were localized to 5 clusters, including the MCL, mPFC, bilateral pITG, and right LOC. Regions that exhibited decreased activity were localized to 3 clusters, including left dlPFC and bilateral aMTG (Threshold-free cluster corrected t-score > 2.3, p < 0.01). **b.** Word cloud of top 5 positive (red) and negative (blue) terms (from 1765) associated with the z-score contrast in activity map (CBP+O > CBP-O). The size of a term is in proportion to the correlation strength of each term to the brain region generated by the Neurosynth reverse inference decoder. **C**. Flowchart depicts the method used to project the whole-brain unthresholded activity contrast map (change in ALFF, CBP+O > CBP-O) into 100 cortical ROIs that were submitted to a multiple regression analysis with the 19 receptor density maps in the same ROI space. **d.** Using a multiple regression analysis, the plot shows the standardized estimates (± 95% CI) for ALFF with receptor density. Spontaneous activity differences (change in ALFF, CBP+O – CBP-O) showed a significant positive relationship with MOR and a negative relationship with serotonin 5HT1A and 5HT1B. **e) S**catter plot for ALFF with MOR (left), 5-HT1A (middle), and 5-HT1B (right). Top brain images show the receptor density maps and bottom brain images show the most positive (red) and negative (blue) regions driving the correlation.

Brain regions that showed increased or decreased ALFF in CBP+O did not statistically significantly correlate with opioid use in CBP+O (**Extended Data Table 12).** This was also the case when comparing ALFF between CBP+O patients on high (MME ≥ 50) versus low (MME <50) doses of opioids (**Extended Data Table 13).** Overall, opioid exposure-related regional activity differences were not statistically significantly related to our clinical or opioid use parameters, suggesting that the activity differences between CBP+O and CBP−O either (1) preexist— perhaps imparting risk for opioid use or confounding by indication, (2) reflect responses to short-term opioid exposure, or (3) adaptations reflecting long-term opioid use. The second and third possibilities can be teased apart by studying CBP+O after they abstain from opioid consumption (see below).

### Cortical neurotransmitter receptor density and its relationship to spontaneous activity in chronic pain patients with long-term opioid use

We studied how receptor density distributions mapped onto CBP+O vs. CBP−O activity differences. Specifically, how much of the cortical ALFF differences between CBP+O and CBP−O can be explained by neurotransmitter receptor densities? To address this issue, we used multiple regression to model the cortical ALFF differences (CBP+O minus CBP−O) using 19 neurotransmitter receptor density maps (constructed by amalgamating PET images from a combined total of 1,238 healthy participants across nine different neurotransmitter systems ^42^) **(Fig 4c).** The receptor density maps explained 51% of the variance of 100 cortical ROIs consisting of the between-group ALFF difference (*p*<0.01)(**Fig 4d**). After adjusting for all of the other receptors, µ-opioid receptor (MOR) density was positively associated with ALFF differences. Conversely, serotonin receptor densities, 5-HT_1A,_ and 5-HT_1B_, were negatively associated with ALFF differences. (**Fig 4e**). The Δ*R*^2^ associated with MOR was 13.7%, 5-HT_1A_ was 19.6%, and 5-HT_1B_ was 17.6%. Together, their Δ*R*^2^ was 27.4%. It is important to note that the regression model remained statistically significant after removing all three primary contributing receptors, indicating while the opioid and serotonin receptors play a significant role, they are not necessarily the sole explanation of ALFF changes in long-term opioid use. These results suggest that the ALFF pattern in CBP+O largely follows specific cortical receptor expression distributions, which may be associated with specific functional characteristics.

### Network dynamics in chronic pain patients with long-term opioid use

We examined how regional activity changes in CBP+O explain the dynamics of resting-state functional networks. We used the eight regional ALFF changes in CBP+O to derive seed-based connectivity matrices (*n=*846 healthy subjects from Connectome1000 ^43^) **(Extended Data Fig. 8).** We derived networks from the seed-based connectivity using PCA; the first PC explained the majority (72%) of the total variance **(Fig. 5a).** After thresholding the first PC (|score| > 0.5), this component was comprised of two opponent networks: an expanded default mode network (DMN) that also incorporated mesocorticolimbic circuits (DMN+MCL), and an expanded task-positive network (eTPN), which included dorsal and ventral attentional and executive control canonical networks ^43^ (**Fig. 5b,c**).

**Fig. 5.**
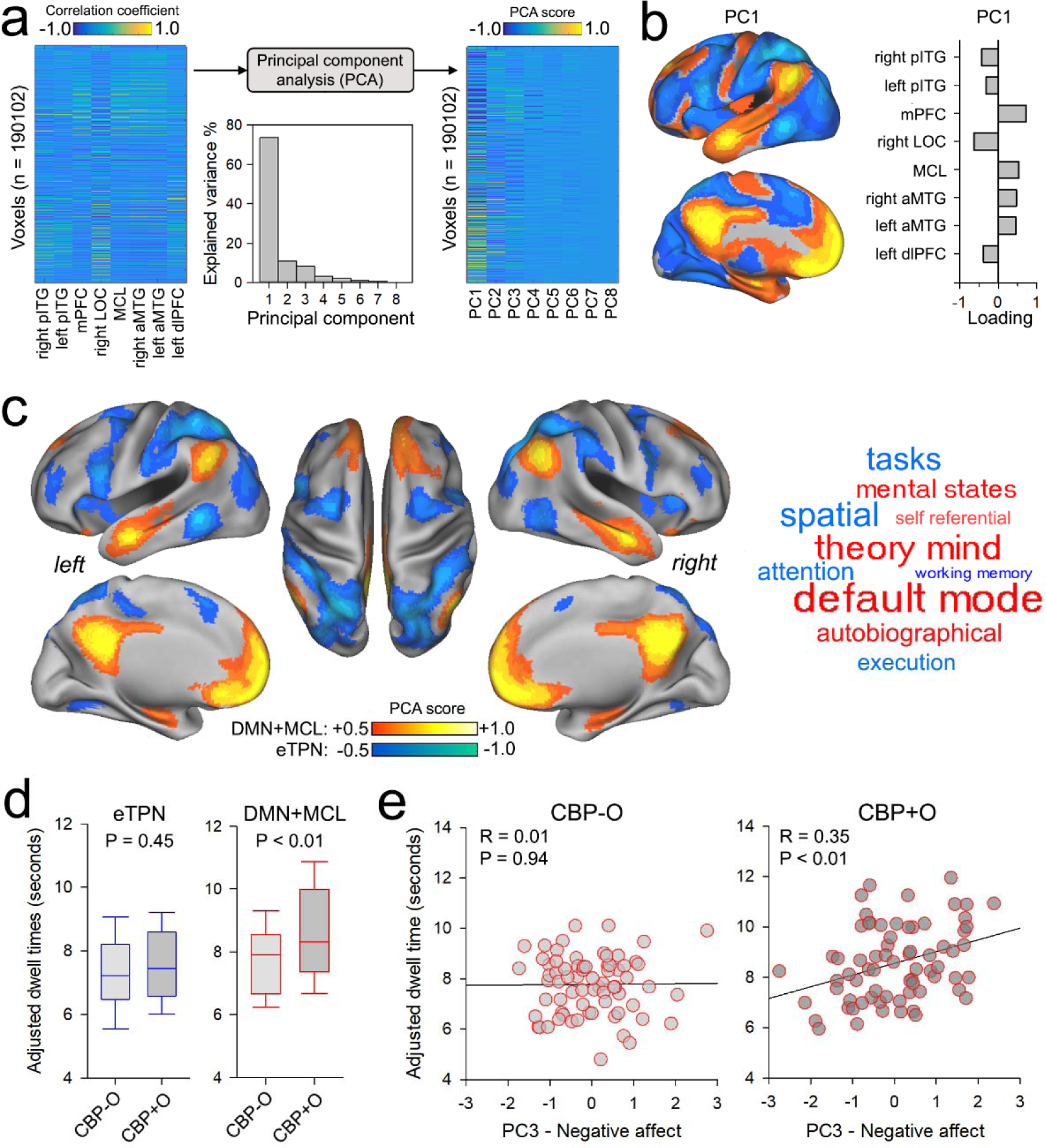
Change in ongoing activity in long-term opioid use is associated with two opponent networks, their dynamics reflecting negative affect only in CBP+O patients. **a.** Whole brain functional connectivity maps of brain regions that showed ALFF differences between CBP+O and CBP-O mapped to one principal component (PC1) that explained 72.34 % of the total variance. **b.** Brain slices show the positive (red-yellow) and negative (blue-green) scores of PC1. Bar graphs display the loading values of functional connectivity maps to PC1. **c**. Brain images show the two brain networks identified from PC1 that included the default mode and the MCL (DMN+MCL) (red-yellow, score > 0.5) and an extended task-positive network (eTPN) that included attention and left and right executive control networks (blue-green, score < -0.5). The word cloud shows the top 5 terms associated with the DMN+MCL network (red) and the eTPN network (blue). The size of a term is in proportion to the correlation strength of each to the corresponding network generated by the Neurosynth reverse inference decoder. **d.** Box plots for dwell times of the DMN+MCL (left) and eTPN (right) networks in CBP-O and CBP+O. The DMN+MCL network showed increased dwell times in CBP+O compared to CBP-O (p<0.01). **e.** The DMN+MCL network dwell times showed a positive association with negative affect in CBP+O but not in CBP-O.

We studied the dynamics of the DMN+MCL and eTPN networks using (1) the dwell times of each network and (2) Pearson correlations between the two networks. Dwell time is how long activity within a network continuously persists (*i.e.*, for how long is the network “on” before turning “off”) ^44^. Compared to CBP−O, CBP+O showed increased dwell time for DMN+MCL (*p*<0.01) but not for eTPN (*p*=0.45) (**Fig. 5d**). Since these networks were derived from eight regions that arose in our ALFF analysis, it seemed plausible that the group difference in network dwell times was attributable to one of the regions. To determine this, we re-estimated the networks’ dwell times after removing the regional BOLD signals from the network BOLD signal. Out of the eight regions tested, only regressing out the MCL rendered dwell times equivalent in duration between CBP+O and CBP−O (**Extended Data Table 14),** demonstrating that increased persistence of DMN+MCL in CBP+O was attributable to MCL activity. In addition to dwell time differences, the DMN+MCL and eTPN were more negatively correlated in CBP+O (*r*=−0.35±0.25) than in CBP−O (*r*=−0.24±0.28; *t*-statistic=2.49, *p*<0.05). Across both groups, these negative correlations were negatively related to DMN+MCL dwell times (*r*=−0.27, *p*<0.05).

We investigated how network dynamics relate to opioid use, COMM, SOWS, and clinical parameters (PC1-3). DMN+MCL dwell times positively correlated with negative affect for CBP+O (*r*=0.34, *p*<0.01), but not CBP−O (*r*=0.01, *p*=0.94) **(Fig. 5e).** Similarly, the correlation between the two DMN+MCL and eTPN was associated with negative affect in CBP+O (*r*=−0.31, *p*<0.01) but not CBP−O (*r*=0.01, *p*=0.96). We validated this finding with an independent negative affect measurement—Negative Affect Summary (NAS) from NIH Toolbox—in a subset of CBP+O (*n*=46) and CBP−O (*n*=22) patients matched for age, pain intensity and duration, and sex (**Extended Data Table 15).** NAS was positively associated with negative affect (PC3) across all 68 patients (*r*=0.74, *p*<0.01) and, consistent with our previous findings^31^, was greater in CBP+O than CBP−O (*t*-statistic = 2.27, *p*<0.05). More importantly, NAS was positively associated with DMN+MCL network dwell times in CBP+O (*r*=0.47, *p*<0.01, n=46) but not in CBP−O (*r*=−0.23, *p* = 0.54, n=22) **(Extended Data Fig. 9).**

### Brief opioid abstinence and network dynamics

To elucidate the relationship between behavior, brain function, and opioid use, in 14 of the CBP+O patients, we perturbed their stable opioid status by requesting the participants to briefly withhold opioid consumption (19.4 ± 6.7 hours). While they abstained, we collected a second resting state fMRI, pain scores (NRS), withdrawal signs (SOWS), and measures of physical and mental well-being (SF12). Blood samples confirmed patients’ abstinence, as there were no or minimal opioids detected in the blood, especially in comparison to their first scan, which was collected within 3.01 ± 2.75 hours of opioid consumption (*p*<0.001) **(Fig 6a, Extended Data Table 16).** Opioid abstinence increased SOWS and decreased SF12 mental scores, but did not statistically significantly change pain intensity and SF12 physical scores (**Fig. 6a, Extended Data Table 16**). Of the regions that showed increased ALFF in CBP+O patients, only the left pITG statistically significantly reduced following abstinence (*p*<0.001). In addition, the right aMTG, a region that shows decreased activity in CBP+O, exhibited an additional large ALFF reduction (**Extended Data Table 17)**. Neither left pITG nor right aMTG ALFF changes exhibited statistically significant correlations with changes in SOWS or SF12 mental scores in CBP+O following abstinence (all *p*>0.2). Moreover, with abstinence, both DMN+MCL and eTPN networks increased their dwell times **(Fig. 6c),** and the changes in DMN+MCL network dwell times correlated with changes in SOWS and in SF12 mental scores. These results show that briefly withholding opioids induces signs of OUD that are associated with network dynamics.

**Fig. 6.**
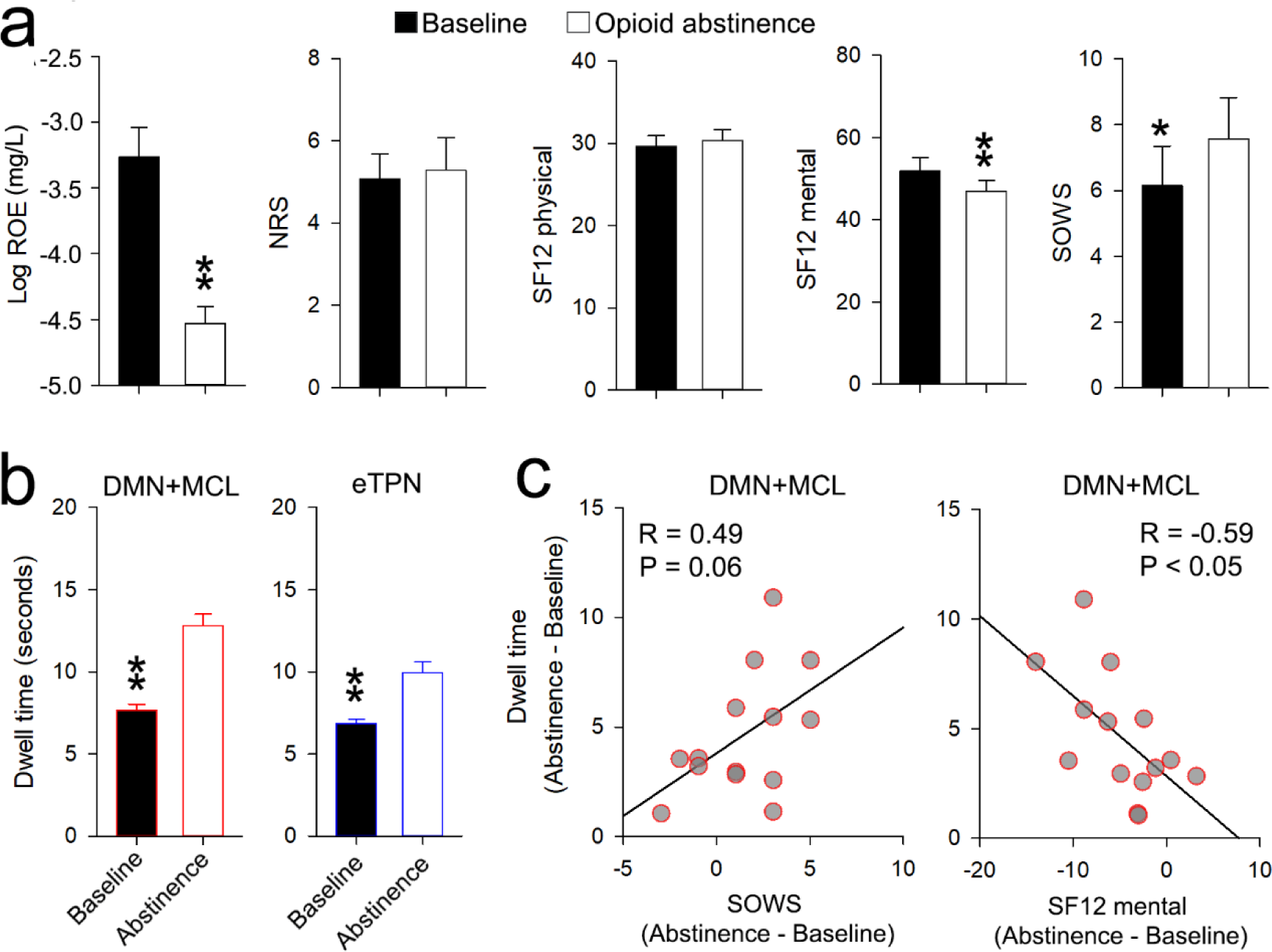
Brief opioid abstinence worsens mental quality and increases withdrawal signs in proportion to increased dwell time for the DMN+MCL network. **a.** Bar graphs represent the mean ± s.e.m of opioid and behavioral measures at baseline (black) and following 19.4 ± 6.7 hours of opioid abstinence (white) in 14 CBP+O patients. Blood levels of opioids (Log ROE), mental well-being (SF12 mental), and withdrawal signs (SOWS) showed significant changes following abstinence. **b.** Dwell times for both the DMN+MCL (red contour) and eTPN (blue contour) networks increased following opioid abstinence. **c.** With opioid abstinence, the change in dwell time of the DMN+MCL network was associated with increased withdrawal signs (SOWS) and decreased mental well-being (SF12 mental). (*p<0.05, **p<0.01, paired t-test). NRS = Numerical Rating Scale; ROE = Relative Opioid Equivalents; SOWS = subjective opioid withdrawal scale.

## Discussion

The present study uncovered how long-term opioid consumption in those with chronic low back pain is associated with specific neurobiological differences. Given that chronic pain and OUD show psychological comorbidities and brain structural and functional maladaptations, the expectation was that CBP+O would substantially deviate from CBP−O—*i.e*., opioid-related adaptations are additive to chronic pain-related adaptations. Behaviorally, long-term opioid use was associated with modestly poorer outcomes than CBP−O. Structurally, opioid use was associated with lower whole-brain gray matter volume and regional gray matter density than CBP−O. These structural changes were also modest compared to those observed in CBP ^35,36^ or substance use disorder (SUD) ^37,38^. Our morphometric results somewhat contradict earlier reports ^45,46^, which show rapid, more extensive, and persistent gray matter decreases; however, these previous reports studied a smaller, opioid-naïve CBP sample. Yet, in line with our data, they also report decreased grey matter volume in mACC.

In contrast to our behavioral and brain morphology findings, we found large differences in ongoing brain activity between CBP+O and CBP−O. The motivational affective (mesocorticolimbic) circuit exhibited the most prominent increased activity in CBP+O, while the dlPFC, which is suggested to provide cognitive control over the mesocorticolimbic pathways ^47^, was the region showing the largest decreased activity. This activity pattern was not related to opioid use, and it was only modestly perturbed following a brief period of opioid abstinence. Thus, we surmise the difference in brain activity between CBP+O and CBP−O is a stable state reflecting long-term adaptations to opioid exposure. Much of this activity pattern could be explained by cortical receptor distributions. There was a strong positive relationship between CBP+O and CBP−O cortical activity differences and MOR expression. As illustrated in Fig. 4e, the mPFC has the highest level of expression of MOR, and this is the same region where ongoing activity increased most in CBP+O relative to CBP-O, while we see the reverse in the visual cortex. There was a similarly strong but negative relationship between 5-HT_1a_ and 5-HT_1b_ receptor expressions and activity difference between CBP+O and CBP−O. Thus, MOR expression seems to drive increased ongoing activity in CBP+O, which maps to the task-negative network (see below), while 5-HT_1a_ and 5-HT_1b_ receptors strongly reflect decreased ongoing activity, mapping them to the task-positive network. We presume these tight receptor-activity relationships are, in part, due to MOR desensitization as a consequence of long-term and regular opioid consumption ^48,49^, which may be accompanied by a compensatory upregulation in cortical 5-HT_1a_ and 5-HT_1b_ receptors.

MORs actions are best studied in the context of OUD and within the mesolimbic VTA, NAc, and mPFC circuitry, where exposure to repeated high doses of opioids leads to tolerance, and following a period of abstinence results in dysphoria and somatic signs of withdrawal ^22^. Much less is known regarding MOR signaling in the cortex with repeated opioid exposure ^50^. Serotonergic neurotransmission is implicated in motivated behaviors in addictive substance-related reward processing ^51^, 5-HT neurons respond to aversive stimuli, and variants of 5-HT_1b_ receptor, including expression in the frontal cortex, are associated with susceptibility to heroin use disorder ^52–54^. The remarkable specificity between activity changes and receptor-type expression levels observed here suggests the utility of different classes of chemicals to control particular parts of the circuitry, for example, serotoninergic antidepressants may aid in opioid tapering. It remains unknown the extent to which the receptor-activity cortical relationship seen here may also exist in OUD or in SUD; such studies remain to be performed.

The relationship between ongoing activity (ALFF) and resting state functional connectivity has been explored in multiple neurological conditions ^55,56^. Yet, to our knowledge, the link between the two has not been studied. Here, using the ALFF activity regional changes as nodes, we identified two opposing functional networks. These networks closely approximate the well-known intrinsic and spontaneously fluctuating task-positive (composed of executive control and attention networks; regions that increase in activity across many tasks) and task-negative (primarily the default mode network, DMN; a circuit that decreases in activity during active tasks) networks, which are strongly anticorrelated in healthy subjects ^57^. CBP+O had a stronger anticorrelation between the two networks and longer task-negative (DMN+MCL) dwell times. The lengthened dwell times were attributable to mesocorticolimbic activity. Following a brief period of opioid abstinence, CBP+O patients had more symptoms of withdrawal, poorer mental well-being, a stronger anticorrelation between the two networks, and greater dwell time in both networks. Together, these results lead to strong conclusions: 1) The mesocorticolimbic circuit seems to be the primary contributor to the network dynamics that underpin abnormal ongoing activity in CBP+O; 2) Since mesocorticolimbic activity controls the dwell time of the task-negative network, long-term adaptations within this circuitry may underlie the enhanced negative affective state of CBP+O. According to the canonical view, the mesocorticolimbic system is responsible for signaling both the reward value and associated expectations. However, recent evidence ^19,58^ and our human fMRI ^59,60^ and rodent studies ^61,62^ implicate this circuit in aversive states and chronic pain. Yet, aversion-related mesocorticolimbic adaptations with long-term opioid exposure remain unknown ^29^.

The CBP+O population studied here included opioid users with no or minimal apparent signs of dependence. Yet, following a brief period of abstinence, they had mild but increased signs of withdrawal and worse mental well-being. Both human and rodent model evidence indicates the causal involvement of the mesocorticolimbic circuit in chronic pain ^11–16^, and this circuit is the main target of opioid exposure leading to OUD ^17,18, 22^. Thus, the change in ongoing activity and network dynamics observed in CBP+O following abstinence should be cast in the context of similar knowledge in OUD and SUD. Changes in ALFF have been reported in OUD studies ^63,64^ but minimally overlap with our results—this may be at least partly attributable to other studies being smaller and having heterogeneous samples (*e.g.*, mixing CBP+O and non-CBP). Multiple lines of task-related fMRI studies in OUD and SUD ^47,65,66^ suggest that the dominant pattern of change in ongoing activity in CBP+O—*i.*e., decreased activity in regions associated with regulating emotion (dlPFC) and increased activity in regions associated with motivated affect (mesocorticolimbic circuits)—is associated with drug craving. However, in CBP+O, this pattern seems to be driven by negative affect; with abstinence, activity and network dynamics are further exacerbated.

Medial and lateral prefrontal cortical activity adaptations are a dominant concept in current models of OUD and SUD ^47,66^, and our results confirm and extend this knowledge to CBP+O. Resting-state functional connectivity differences have been reported for OUD by multiple groups, primarily for mesocorticolimbic subregions, see ^15^. Some of these results contradict each other, which, again, could be attributable to small and heterogeneous samples. Network dynamics and dwell time were recently studied in those with OUD and alcohol use disorder (AUD) in comparison to healthy subjects ^67^. The authors observed shorter dwell times in OUD and AUD relative to healthy controls for the DMN-dominated task-negative networks. This is opposite to our DMN+MCL results, perhaps for multiple reasons: (1) network expanse between the two studies (including the mesocorticolimbic system in our case)—however, even after removing the influence of the mesocorticolimbic circuits, our results are still inconsistent with those of ^67^; (2) CBP+O’s opioid use is overseen by physicians and they may not have OUD; and (3) opioids may not have the same additive effect in CBP as in non-CBP subjects, as CBP−O’s brain networks are already different from healthy individuals ^11^. Overall, activity and network dynamics in CBP+O share properties with OUD and SUD, especially concerning their pattern, yet, dwell times may diverge between CBP+O and SUD.

## Conclusions

In the first large-scale assessment of the psychobiology of stable and long-term opioid use in CBP, in comparison to a closely matched CBP not using opioids, we found only modestly worse psychological phenotypes and small brain structural decreases. However, CBP+O also showed large changes in spontaneous brain activity, dominated by increased mesocorticolimbic and decreased dlPFC activity. These activity changes mapped onto two competing networks, the dynamics of which correlated with negative affect and signs of withdrawal following opioid abstinence. The strong and circuit-specific relationships between MOR, 5-HT_1a_, and 5-HT_1b_ receptor expressions with changes in spontaneous activity in CBP+O users suggest novel receptor-specific treatment options, especially for opioid tapering.

## Contributions

A.V.A designed and oversaw all aspects of the study. M.N.B, G.R., A.D.V., R.J. and P.B. performed all data analysis. T.J.S., O.C. and G.R. recruited patients and collected pain and behavioiral questionnaires. L.J., G.R. and R.J. collected and organized brain imaging data. J.G. and A.D.W. provided clinical input. M.N.B. and A.V.A. drafted the manuscript, with contributions from all authors.

## Data Availability

All data produced in the present study are available upon reasonable request to the authors.

## Acknowledgments

We would like to thank all the patients who participated in this study. We gratefully acknowledge the discussions and suggestions by all members of the Center for Translational Pain Research, and especially the project leaders, namely Drs. R. Awatramani, T. Lerner, M. Martina, and D. Surmeier. The study was supported by the Center of Excellence for Chronic Pain and Drug Use at Northwestern University NIH NIDA P50DA044121 (PI A.V.A.), and in part by DOD grant W81XWH-17-1-0426 (PI T.J.S.).

## Notes

### Competing Interest Statement

The authors have declared no competing interest.

### Funding Statement

This study was funded by the National Institute of Drug Abuse of the National Institutes of Health Grant No. 1P50DA044121, and the
National Science Foundation Graduate Research Fellowship under Grant No. DGE-132458, and by the National Institute Of
Neurological Disorders And Stroke of the National Institutes of Health under Award No. F31NS126012.

### Author Declarations

Northwestern University Institutional Review Board.

