## Supplementary material for "Neuropsychology of chronic back pain managed with long-term opioid use": Methods

### Online methods

### Participants.

### We recruited 70 CBP patients on opioid therapy for at least 6 months (CBP+O) and 70 patients not taking opioids (CBP-O) one-to-one matched for age, sex, pain intensity, and pain duration (from a pool of 150 CBP-O). All participants are over 18 years old, fluent in English, and understand instructions and questionnaires. Recruitment was from the Northwestern Medicine (NM) healthcare system and Shirley Ryan Ability Lab. The exclusion criteria included (1) treatment with a spinal cord stimulator; (2) diagnosis of rheumatoid arthritis, ankylosing spondylitis, acute vertebral fractures, fibromyalgia, low back/spine oncologic history, and other comorbid neurological disorders, including major depression and psychiatric disorder requiring treatment; (3) involvement in litigation regarding their back pain, having a disability claim, or receiving workman’s compensation; (4) significant comorbid diseases such as uncontrolled hypertension, unstable diabetes mellitus, renal insufficiency, congestive heart failure, coronary or peripheral vascular disease, chronic obstructive lung disease, or malignancy; (5) Pregnancy during the study. This project was approved by the Northwestern Institutional Review Board (STU00207384, STU00205398).

**Demographic and general health measurements**

All participants completed a personal health history questionnaire, including information about age, gender, race, ethnicity, height, weight, alcohol use, and smoking status.

**Pain and behavioral measurements**

Pain duration and pain characteristics were recorded for all participants. Pain characteristics were assessed using the Numerical Rating Scale NRS, McGill Pain Questionnaire – Short Form (sf-MPQ), and Pain Detect. The NRS is an 11-point numerical rating scale used to measure pain intensity, with “0” corresponding to no pain and “10” to the worst pain possible (or imaginable) (1). The sf-MPQ is a 15-item measure that separates the sensory and affective components of pain (2). Finally, the PainDETECT consists of seven items assessing the neuropathic pain (3). We also collected the Pain Catastrophizing Scale (PCS), a survey with 13 items asking about thoughts or feelings related to pain experiences (4).

Mood and emotional measures included the Beck Depression Inventory (BDI**),** a 21 multiple choice self-reported questionnaire evaluating the severity of depression (5), the Positive and Negative Affect (PANAS): 20 items self-report questionnaire measuring positive and negative affect (6), and the Pain Anxiety Symptoms Scale (PASS): 20 questions measuring chronic pain related anxiety and fear (7).

We also collected the Patient-Reported Outcomes Measurement Information System (PROMIS), An 57 item questionnaire evaluating important health-related quality of life domains assessing the seven domains of pain interference, fatigue, depression and sadness, anxiety and fear, sleep disturbance, physical function, and social activity (8), as well as the Short Form Health Survey (SF-12), a self-reported questionnaire to evaluate the impact of health on one’s quality of life. It results in two physical and mental health scores (9). Finally, disability was assessed using Oswestry Low Back Pain Disability (ODI), a ten-item questionnaire measuring functional disability in activities of daily living in patients with low back pain (10).

**Multidimensionality of pain and behavioral measures**

The internal structure and multidimensionality of pain and behavioral measures were assessed using principal component analysis (PCA) in Matlab (R2020a, Statistics and Machine Learning Toolbox Version 11.7) with varimax rotations across all patients. Principal components with eigenvalue > 1 that explained > 10% of variance were retained, resulting in three components (**Extended Data Table 2**). To ensure that the components were not related to opioid use, we also performed PCA for CBP+O and CBP-O separately. The similarity between the principal components determined from different groups was assessed by comparing their corresponding loading factors using correlation analysis **(Extended Data Fig 1).**

#### Opioid use measures

#### For the group using opioids, we collected details of their opioid prescription. We converted the daily dosage to morphine milligram equivalent (MME) using the standard conversion factors generated by the CDC (11). The MME is used to estimate the dose of morphine based on the dose of the opioid, route of administration, and conversion ratio. In addition, we collected the duration of opioid use (DOU) for each patient. We also collected blood samples from participants immediately after brain scans to quantify systemic levels of opioids. The concentrations of eight analytes (oxymorphone, hydromorphone, oxycodone, hydrocodone, fentanyl, buprenorphine, methadone, and tramadol), and the presence of three others (morphine, codeine, and heroin) in plasma were determined by liquid chromatography-tandem mass spectrometry after sample preparation by solid-phase extraction, using deuterated analogs (oxymorphone-D3, hydromorphone-D6, oxycodone-D6, hydrocodone-D6, buprenorphine-D4, methadone-D3, and tramadol-C13D3) as internal standards, based on the method reported by Langman et al (12). One hundred microliters of the plasma sample were diluted with 300 uL of 1% acetic acid, vortexed, and loaded onto a Phenomenex Strata-X-Drug B 96-well plate. After washing with 100 mM sodium acetate buffer and methanol, the analytes were eluted with an ethyl acetate/isopropanol/ammonium hydroxide (7:2:1 v/v/v) mixture and evaporated to dryness under a stream of nitrogen. The samples were reconstituted in 15% methanol in water and analyzed by a Sciex 6500+ Qtrap LC-MS/MS system equipped with a Shimadzu Nexera2 UPLC system. Samples were eluted from a Phenomenex Kinetex 2.6 μm Biphenyl (50 x 2.1 mm) column with a mobile phase consisting of A: water with 0.1 % formic acid and B: methanol with 0.1 % formic acid at a flow rate of 0.6 mL/min with gradient from 10% B to 85% B in 4.5 min. Samples were run in duplicate, and the average opioid concentration was used. The various opioid concentrations mg/L were then converted to a relative opiate equivalent (ROE), taking into consideration the affinity of each opioid to the mu-opioid receptor (MOR) (13) and the respective molar weights (14-21). We realize that limiting tramadol to its MOR affinity excludes other aspects of the tramadol effect; however, we are specifically interested in the opiate activity on MOR. Samples with peak presence of opioids below the lower limit of quantitation were imputed as the halfway point from zero to the measurable lower quantifiable threshold. The MME and ROE were log-transformed and compared to the characteristics of pain and psychosocial measures.

**Opioid behavior measures**

We also evaluated the risk of opioid misuse measured with the Current Opioid Misuse Measure (COMM) and the intensity of opioid withdrawal using The Subjective Opioid Withdrawal Scale (SOWS). The COMM is a 17 self-reported item questionnaire measuring aberrant medication-related behavior among persons with chronic pain who are prescribed opioids for pain (22). The SOWS measures the intensity of opioid withdrawal using 16 symptoms that the subjects rate in intensity (23).

**MRI acquisition**

All participants were scanned with a 3 Tesla Siemens Magnetom Prisma whole-body scanner using a 64 channel-head/neck coil. T1-anatomical brain images were acquired using the following parameters: A voxel size of 1 × 1× 1mm^3^, a repetition time/echo time (TR/TE) = 2.3s/2.4ms, a flip angle of 9°, the in-plane resolution = 256 × 256, using 176 slices per volume and a field of view of 256mm. Resting-state fMRI images were acquired using the following parameters: A voxel size of 2 × 2 × 2mm^3^, a TR/TE = 555ms/22ms, a flip angle of 47°, the in-plane resolution = 96 × 104, using 1110 volumes, a multiband accelerator = 8 and 64 slices acquired with interleaved ordering, covering the entire brain from the cerebellum to the vertex. Diffusion tensor images were acquired for only 58/70 participants in each group. These were acquired with a two-shell dMRI protocol: the first acquisition had 64 directions and a bval = 2000, and the second with 30 directions and a bval = 700. Both had a voxel size of 2 × 2 × 2mm^3^, a TR/TE = 3.5s/ 9.2ms, and a flip angle 90, the in-plane resolution = 116 × 116 and 72 interleaved slices.

**fMRI preprocessing**

Preprocessing of resting-state fMRI images was performed using the standardized FMRIPREP pipeline. Following the removal of the first 100 volumes to eliminate saturation effects and achieve steady-state magnetization, the following steps were performed: motion correction, intensity normalization, nuisance regression of 6 motion vectors, signal-averaged overall voxels of the eroded white matter and ventricle region, and global signal of the whole brain. Following this, to obtain the low-frequency fluctuations of the resting-state fMRI signal, the BOLD time series were band-pass filtered to 0.008Hz – 0.1Hz by applying a Butterworth filter. All pre-processed fMRI data was registered to the MNI152 2mm template by using a two-step procedure, in which the mean of the preprocessed fMRI data was registered with a 7-degrees-of-freedom affine transformation to its corresponding T1 brain (FLIRT); transformation parameters were computed by nonlinearly registering individual T1 brains to the MNI152 template (FNIRT). Combining the two transformations by multiplying the matrices yielded transformation parameters normalizing the pre-processed fMRI data to the standard space.

**Amplitude of low-frequency fluctuations (ALFF)**

##### ALFF analysis (24) was performed on preprocessed data using Matlab software. The time series for each voxel was transformed to the frequency domain, and the power spectrum was then obtained. Since the power of a given frequency is proportional to the square of the amplitude of this frequency component, the square root was calculated at each frequency of the power spectrum, and the averaged square root was obtained across 0.01–0.08 Hz at each voxel. This averaged square root was taken as the ALFF and divided by the individual global mean of ALFF within a brain mask, obtained by removing the tissues outside the brain. This standardization procedure is like that used in PET studies. Finally, we conducted spatial smoothing using an isotropic Gaussian kernel of 5 mm full width at half-maximum. Differences in ALFF between groups (CBP+O > CBP-O) were performed using randomize in FSL. Gray matter density and temporal signal-to-noise ratio were used as voxel-wise regressors, while age, MQS, sex, race, pain intensity, pain duration (log), and intracranial volume were used as group-wise regressors. Correction for multiple comparisons was performed using the Threshold-Free Cluster Enhancement (TFCE) method (FWE *p* < 0.01).

**Region of the interest analysis**

Regions of interest (ROI) that showed significant ALFF changes between CBP+O and CBP-O were determined post-hoc from the whole-brain ALFF (CBP+O > CBP-O) contrast map using Easythresh, a simple clustering algorithm in FSL that identifies spatially contiguous clusters. We identified five clusters (ROIs) that showed increased ALFF (FEW p <0.01) in CBP+O compared to CBP-O and three clusters that showed decreased ALFF in CBP+O (FEW p <0.01) compared to CBP-O.

### Peripheral gray matter volume (PGMV) analysis

### Global brain volume for each subject was estimated using SIENAX in FSL. It is an automated process that utilizes FSL programs to strip the non-brain tissue from the estimated peripheral gray matter volume (PGMV). The difference in PGMV between CBP+O and CBP-O was determined using ANCOVA analysis with age, sex, race, pain intensity, pain duration (log), BMI, MQS, and intracranial brain volume as regressors. The relationship between PGMV and behavioral or opioid measures was examined using Pearson correlation analysis after adjusting for covariates of no interest.

##### Voxel-based morphometry (VBM) analysis

##### Gray matter density (GMD) was examined using voxel-based morphometry from FSL-VBM. All T1-weighted images were first brain extracted and then segmented into gray matter, white matter, or cerebrospinal fluid. A common gray matter template was generated for patients by registering and averaging all gray matter images. The gray matter image of each participant was then registered to the common template using non-linear transformation. Differences between groups (CBP+O > CBP-O) were performed using randomize in FSL, with age, MQS, sex, race, pain intensity, pain duration (log), and intracranial volume as regressors. Correction for multiple comparisons was performed using the Threshold-Free Cluster Enhancement (TFCE) method (FWE *p* < 0.01).

##### Subcortical volume analysis

### Volumes of subcortical regions were obtained using FSL. More detailed information about the processing steps of subcortical segmentation by FSL-FIRST can be found in Patenaude et al. (2011). In this study, we segmented the T1 images using FSL-FIRST with none boundary correction. Volumes of subcortical regions were determined using a voxel count. Differences in subcortical volumes between CBP+O and CBP-O were determined using ANCOVA analysis with age, sex, race, pain intensity, pain duration (log), BMI, MQS, and intracranial brain volume as regressors.

##### Diffusion tensor images (DTI) analysis.

##### Fractional anisotropy (FA) maps were used as a proxy for white-matter integrity. DTI analyses were conducted using tools from the FMRIB diffusion toolbox (FDT). First, diffusion-weighted images were visually inspected for gross artifacts. No visible quality control issues were identified. Images were then corrected for eddy current distortions and head movement using EDDY(25). For additional unbiased quality control, we used QUAD and SQUAD (26). No subjects were classified as outliers, given quality control metrics. DTIFIT was used to fit a tensor to the data, calculate principal directions, and extract fractional anisotropy maps. FA images were analyzed with Tract-Based Spatial Statistics Field (27) (TBSS), which provides good inter-subject alignment for white-mater images to perform group-level analyses. Diffusion images were transformed into standard space (FMRIB58) through non-linear transformation and then projected to the mean skeletonized images, thresholded at the default value of FA = 0.2. Statistical analyses were conducted using the same design and methods as VBM analyses plus an additional motion confound as a covariate of no interest (relative motion estimated by QUAD). Statistical analyses were also conducted using Randomize with TFCE cluster enhancement (optimized for TBSS analyses, option -T2) and FWE correction for multiple comparisons.

**Spatial association between ALFF and receptor density maps**

The un-thresholded whole-brain contrast map ALFF (CBP+O > CBP-O) was parcellated to 100 cortical brain regions according to the Schaefer atlas (27). The relationship of the regional ALFF z-scores with the corresponding z-scores of 19 different neurotransmitter receptors and transporters density maps obtained from Hansen et al. (28) was determined using a multiple regression analysis with the ALFF z-scores as the dependent variable and the z-scores from 19 receptors as the independent variables.

##### ROI-based functional connectivity networks.

##### We used seed-based analysis to generate whole-brain functional connectivity maps for the eight ROIs that showed ALFF differences between CBP+O and CBP-O in 836 healthy subjects obtained from Connectome-1000. For each subject, the functional connectivity map for each ROI was determined by correlating the mean bold signal of the ROI with the bold signal of all voxels in the brain. The mean brain functional correlation map for each ROI was then determined by averaging the maps for any given ROI across all subjects (Extended data Fig 8). The spatial similarities and dependencies of the eight functional correlated networks were assessed using PCA in Matlab (R2020a, Statistics and Machine Learning Toolbox Version 11.7). All maps projected to one component that was used to determine two networks: 1) DMN+MCL, identified by thresholding the PCA scores > 0.5, and 2) the eTPN, identified by thresholding the PCA score <-0.5.

##### Dwell times

##### Overall bold signal for DMN+MCL and eTPN was determined using the weighted mean from all voxels within the network. For each patient, the dwell time for each network was calculated as the mean duration in seconds of temporally continuous runs where bold was in active state (bold > mean bold). Differences in Dwell times between CBP+O and CBP-O were determined using ANCOVA analysis with age, sex, race, pain intensity, pain duration (log), BMI, MQS, and intracranial brain volume as regressors of no interest. The relationship between dwell times and behavioral or opioid-related measures was examined using Pearson correlation analysis after adjusting for covariates of no interest. Differences in dwell times following abstinence were determined using a paired t-test.
