## Extended data for "Neuropsychology of chronic back pain managed with long-term opioid use"

^1^ Center for Translational Pain Research, ^2^ Department of Neuroscience, ^3^ Department of Anesthesia, ^4^ Physical Medicine and Rehabilitation, ^5^ Biomedical Engineering and Statistics & Data Science, ^6^ Medical and Social Sciences, Northwestern University, Chicago, Illinois; ^8^ Shirley Ryan Ability Lab, Chicago, Illinois. ^9^ Department of Anesthesiology and Perioperative Medicine, School of Medicine, University of Pittsburgh, Pittsburgh, Pennsylvania.

**Extended Tables 1-17, figures 1-9.**

|  | **CBP-O (n=70)**  mean ± s.d. | **CBP+O (n=70)**  mean ± s.d. | **p-value** |
| --- | --- | --- | --- |
| ***Demographic and general health characteristics*** | | | |
| Age (years) | 59.87 ± 11.97 | 60.10 ± 10.50 | 0.87 |
| Sex/female (%) | 46 (65.71%) | 46 (65.71%) | 1 |
| Alcohol (%) | 46 (65.71%) | 33 (47.14%) | 0.06 |
| Smoking (%) | 13 (18.57%) | 15 (21.42%) | 0.90 |
| Race  White | 41 (58.57%) | 45 (64.29%) | 0.47 |
| Black | 15 (21.43%) | 23(32.86%) | 0.23 |
| Other/Undisclosed | 14 (20.00%) | 2 (2.86%) | 0.01 |
| BMI | 30.12 ± 5.21 | 31.16 ± 7.36 | 0.35 |
| MQS | 10.33 ± 7.56 | 20.53 ± 12.35 | <10^-5^ |
| ***Pain characteristics*** | | | |
| Pain intensity (NRS) | 5.34 ± 2.13 | 5.40 ± 1.94 | 0.87 |
| Pain duration (years) | 16.64 ± 19.16 | 17.21± 12.36 | 0.68 |
| ***Opioid consumption and behavior*** |  |  |  |
| MME | -- | 41.41 ± 70.06 |  |
| ROE (mg/L) | -- | 0.0042 ± 0.009 |  |
| Duration of opioid use (years) | -- | 7.80 ± 6.09 |  |
| SOWS |  | 8.83 ± 8.27 |  |
| COMM | -- | 7.17 ± 6.10 |  |

**Extended data Table 1. Patient demographics and general health.**

CBP+O and CBP-O patients were matched one-to-one between 70 CBP+O and a select 70 of 150 CBP-O, for age, sex, alcohol use, cigarette smoking, and BMI (two-tailed unpaired *t*-tests or chi-square tests were performed). They were also matched for pain intensity and duration.

Opioid consumption was quantified using the daily prescription converted into MME, the blood levels of opioids converted to an ROE (mg/L), and the duration of opioid use in years. On average, CBP+O exhibited low opioid use (MME < 50) and high non-opioid medication use compared to CBP-O. On average, opioid blood concentrations were 10 times lower than those usually observed in heroin misuse (around 0.05 mg/L) and more than 25 times less than those detected in heroin overdose death (>0.1 mg/L). The CBP+O patients with the highest opioid blood level displayed an ROE value of 0.048 mg/L, which corresponded to the second-highest MME value of 210. The overall low opioid use in our patients was consistent with the observed low withdrawal symptoms (SOWS) and misuse symptoms (COMM).

There was no relationship between pain duration and duration of opioid use in CBP+O. Data presented as mean ± s.d.. BMI = body mass index; MQS = medication quantification scale. NRS = numerical rating scale; MME = daily morphine milligram equivalent; ROE = relative opioid equivalent. SOWS = subjective opioid withdrawal scale; COMM = current opioid misuse measurement.

|  | **CBP (n=140)** | | | **CBP-O (n=70)** | | | **CBP+O (n=70)** | | |
| --- | --- | --- | --- | --- | --- | --- | --- | --- | --- |
|  | **PC1** | **PC2** | **PC3** | **PC1’** | **PC2’** | **PC3’** | **PC1’’** | **PC2’’** | **PC3’’** |
| PainDETECT | 0.22 | **0.77** | 0.08 | 0.13 | **0.82** | 0.15 | 0.31 | **0.70** | 0.07 |
| MPQ sensory | 0.16 | **0.88** | -0.01 | 0.05 | **0.91** | -0.05 | 0.23 | **0.81** | 0.05 |
| MPQ affect | -0.01 | **0.82** | 0.15 | -0.07 | **0.70** | 0.09 | 0.01 | **0.88** | 0.17 |
| PCS | 0.28 | 0.42 | **0.53** | 0.44 | 0.23 | **0.59** | 0.17 | **0.61** | 0.47 |
| ODI | **0.73** | 0.43 | 0.21 | **0.76** | 0.32 | 0.14 | **0.62** | 0.51 | 0.26 |
| PROMIS function | **-0.86** | -0.07 | -0.17 | **-0.84** | 0.19 | -0.04 | **-0.83** | -0.23 | -0.21 |
| PROMIS anxiety | 0.21 | 0.01 | **0.74** | 0.34 | -0.01 | **0.64** | 0.18 | 0.08 | **0.78** |
| PROMIS depression | 0.20 | 0.04 | **0.80** | 0.25 | -0.05 | **0.73** | 0.19 | 0.13 | **0.82** |
| PROMIS fatigue | **0.60** | 0.08 | 0.51 | **0.55** | -0.01 | 0.50 | **0.61** | 0.06 | 0.52 |
| PROMIS sleep dis | 0.42 | 0.05 | 0.49 | **0.53** | 0.06 | 0.49 | 0.44 | 0.01 | 0.48 |
| PROMIS social | **-0.81** | -0.06 | -0.38 | **-0.81** | 0.11 | -0.36 | **-0.78** | -0.16 | -0.35 |
| PROMIS pain Int. | **0.82** | 0.08 | 0.32 | **0.85** | -0.11 | 0.26 | **0.74** | 0.17 | 0.32 |
| SF12 physical | **-0.88** | -0.18 | -0.04 | **-0.88** | -0.14 | -0.03 | **-0.84** | -0.18 | -0.01 |
| SF12 mental | -0.05 | -0.26 | **-0.79** | 0.07 | 0.03 | **-0.81** | -0.08 | -0.34 | **-0.78** |
| BDI | 0.39 | 0.23 | **0.68** | 0.25 | 0.03 | **0.77** | 0.41 | 0.20 | **0.70** |
| PANAS positive | -0.18 | 0.15 | **-0.61** | -0.16 | 0.43 | **-0.43** | -0.19 | -0.02 | **-0.68** |
| PANAS negative | 0.11 | 0.45 | **0.67** | -0.01 | 0.36 | **0.70** | 0.20 | 0.45 | **0.67** |
| Eigen value | 4.34 | 2.80 | 4.19 | 4.52 | 2.55 | 3.95 | 4.01 | 3.10 | 4.39 |
| Prob Exp Variance | 0.26 | 0.16 | 0.25 | 0.27 | 0.15 | 0.23 | 0.24 | 0.18 | 0.26 |

**Extended data Table 2. Principal component analysis for all participants (CBP, n = 140), or when performed separately for each subgroup, CBP-O, and CBP+O.**

Factor loadings were very similar for all three analyses.

The table displays the loading of all seventeen measurements on the principal components determined from all patients (PC1, PC2, PC3), CBP-O (PC1’, PC2’, PC3’) and CBP+O (PC1’’, PC2’’, PC3’’). Data from all patients, or separately for CBP+O and CBP-O, mapped to the same three principal components that represent functional disability (PC1), pain quality (PC2), and negative affect (PC3).

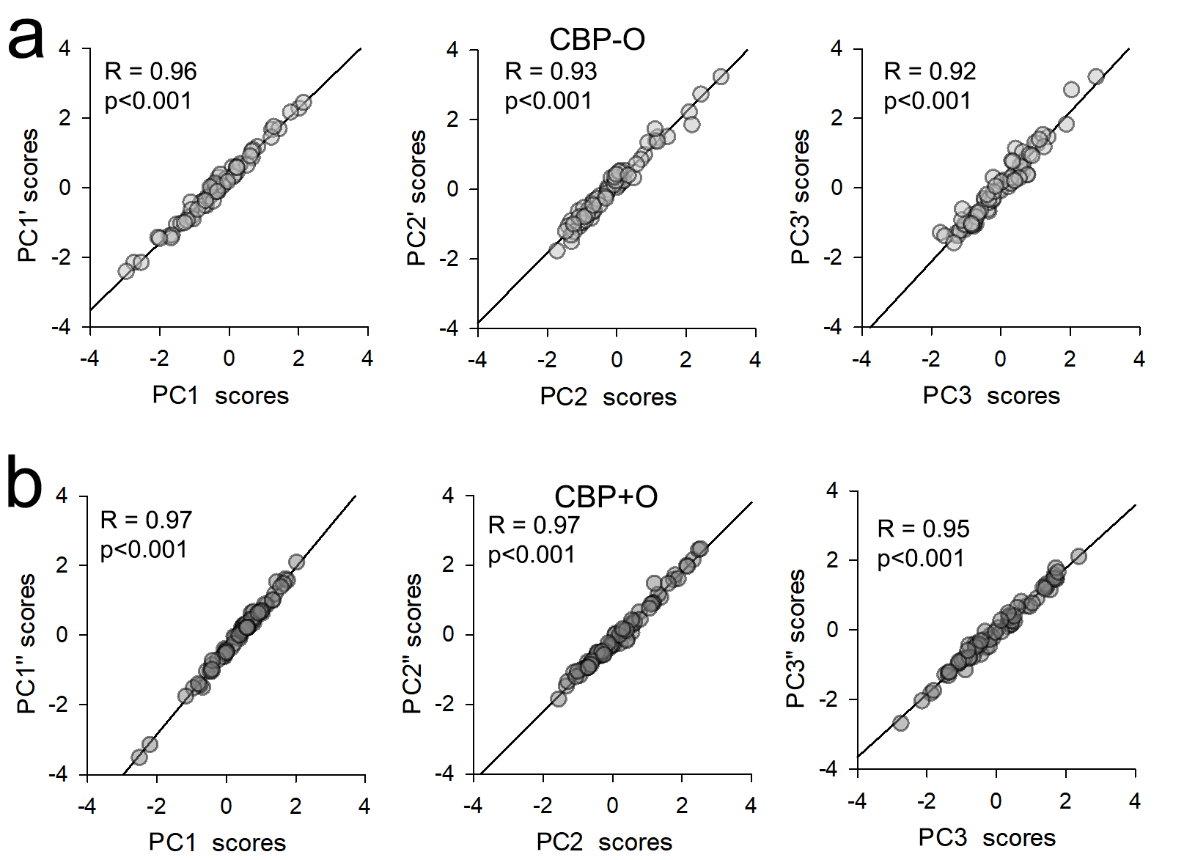

**Extended Data Fig. 1. Behavioral and clinical properties of CBP+O and CBP-O patients exhibited similar principal components decomposition.**

**a.** Scatter plots show the relationship of principal components scores determined from CBP-O (PC1’, PC2’, PC3’) with component scores computed from all patients (PC1, PC2, PC3). **b.** Scatter plots show the relationship of principal component scores determined from CBP+O (PC1’’, PC2’’, PC3’’) with component scores computed from all patients (PC1, PC2, PC3).

Overall, principal components computed from CBP-O and CBP+O showed similar scores to those computed from all patients (all R-values > 0.92; p<0.01).

|  | **PC1 (Functional disability)** | | | | **PC2 (Pain Quality)** | | | | **PC3 (Negative affect)** | | | |
| --- | --- | --- | --- | --- | --- | --- | --- | --- | --- | --- | --- | --- |
|  | **Beta** | **η_p_^2^** | **F-value** | **P-value** | **Beta** | **η_p_^2^** | **F-value** | **P-value** | **Beta** | **η_p_^2^** | **F-value** | **P-value** |
| ***Opioid (CBP-O > CBP+O)*** | **-0.369** | **0.115** | **12.71** | **<10^-3^** | **-0.305** | **0.078** | **8.29** | **<10^-2^** | 0.060 | 0.003 | 0.26 | 0.61 |
| ***Sex (Male > Female)*** | -0.014 | 0.000 | 0.03 | 0.87 | -0.057 | 0.004 | 0.39 | 0.54 | 0.107 | 0.011 | 1.14 | 0.29 |
| ***Race (White > Black)*** | -0.007 | 0.000 | 0.01 | 0.94 | -0.015 | 0.000 | 0.03 | 0.87 | 0.081 | 0.006 | 0.64 | 0.43 |
| ***Age*** | 0.187 | 0.039 | 3.93 | 0.06 | -0.064 | 0.004 | 0.41 | 0.56 | **-0.236** | **0.048** | **4.99** | **0.03** |
| ***BMI*** | **0.247** | **0.068** | **7.16** | **0.01** | -0.007 | 0.000 | 0.00 | 0.94 | -0.124 | 0.014 | 1.42 | 0.24 |
| ***NRS*** | 0.148 | 0.026 | 2.62 | 0.11 | **0.372** | **0.140** | **15.90** | **<10^-3^** | -0.042 | 0.002 | 0.16 | 0.69 |
| ***Log Pain duration*** | -0.118 | 0.017 | 1.65 | 0.20 | 0.006 | 0.000 | 0.00 | 0.95 | -0.002 | 0.000 | 0.00 | 0.99 |
| ***MQS*** | 0.070 | 0.005 | 0.46 | 0.50 | -0.033 | 0.001 | 0.10 | 0.75 | 0.089 | 0.006 | 0.60 | 0.44 |

**Extended Data Table 3. Analysis of covariance results for principal components with opioid use (CBP-O > CBP+O), demographics (sex, race, age, BMI), and pain parameters (NRS, Log Pain Duration, MQS).**

Opioid use exacerbated functional disability (PC1) and pain quality (PC2).

We also observe that PC1 is worsened with higher BMI, PC2 worsened with higher pain scores, and PC3 improved with age. These additional relationships are consistent with the labels we assign to the three PCs.

Effect size and significance of opioid use, demographic and pain properties on clinical and behavioral components of CBP patients. Significance was determined using ANCOVA. Effect size was computed using partial eta-squared (**η_p_^2^**). **η_p_^2^**<0.05 indicates small effect size; 0.05< **η_p_^2^**B<0.15 indicates medium effect size; **η_p_^2^**> 0.15 indicates large effect size indicate small effect size BMI = body mass index; MQS = medication quantification scale. NRS = numerical rating scale.

|  | **CBP-O (n=70)**  mean ± s.d. | **CBP+O (n=70)**  mean ± s.d. | **t-score (p-value)** |
| --- | --- | --- | --- |
| ***PainDETECT*** | 11.78 ± 7.56 | 15.06 ± 7.53 | -2.57 (0.01) |
| ***MPQ sensory*** | 11.48 ± 6.84 | 14.87 ± 6.75 | -2.94 (<10^-2^) |
| ***MPQ affect*** | 2.89 ± 2.67 | 3.81 ± 3.33 | -1.82 (0.07) |
| ***PCS*** | 13.27 ± 10.09 | 14.53 ± 11.50 | -0.68 (0.49) |
| ***ODI*** | **30.22 ± 13.02** | **41.42 ± 13.17** | **-5.06 (<10^-5^) *** |
| ***PROMIS function*** | **41.12 ± 6.51** | **36.59 ± 5.72** | **4.35 (<10^-5^) *** |
| ***PROMIS anxiety*** | 50.71 ± 8.77 | 50.72 ± 8.89 | -0.01 (0.99) |
| ***PROMIS depression*** | 47.64 ± 7.89 | 48.93 ± 8.52 | -0.94 (0.35) |
| ***PROMIS fatigue*** | **51.44 ± 8.22** | **56.69 ± 8.30** | **-3.76 (<10^-3^) *** |
| ***PROMIS sleep dis*** | 53.21 ± 8.21 | 54.26 ± 10.35 | -0.67 (0.51) |
| ***PROMIS social act*** | **49.15 ± 8.70** | **43.82 ± 7.80** | **4.04 (<10^-4^) *** |
| ***PROMIS pain Int.*** | **58.98 ± 6.26** | **63.19 ± 5.61** | **-4.18 (<10^-4^) *** |
| ***SF12 physical*** | **37.93 ± 9.74** | **31.43 ± 8.49** | **4.21 (<10^-4^) *** |
| ***SF12 mental*** | 52.49 ± 8.57 | 48.46 ± 11.12 | 2.40 (0.02) |
| ***BDI*** | **5.97 ± 4.51** | **11.03 ± 7.76** | **-4.69 (<10^-5^) *** |
| ***PANAS positive*** | 32.28 ± 7.76 | 31.30 ± 7.91 | 0.73 (0.46) |
| ***PANAS negative*** | 16.52 ± 5.11 | 18.59 ± 6.08 | -2.17 (0.03) |

**Extended data Table 4. Differences in seventeen pain and mood scores between CBP+O and CBP-O patients.**

In CBP+O as compared to CBP-O, all pain and mood measures tended towards or showed worse outcomes, and none indicated improvement.

CBP+O and CBP-O patients showed significant differences across multiple emotional, functional and pain properties (two-tailed t-test, *p<0.05 Bonferroni corrected. MPQ = McGill Pain Questionnaire; PCS = Pain Catastrophizing Scale; ODI = Oswestry low back Disability Index; PROMIS = Patient-Reported Outcomes Measurement Information System, subscales are labeled, pain int. = pain interference; SF12 = Short Form quality of life scale; BDI = Becks Depression Inventory; PANAS = Positive and Negative Affect Schedule.

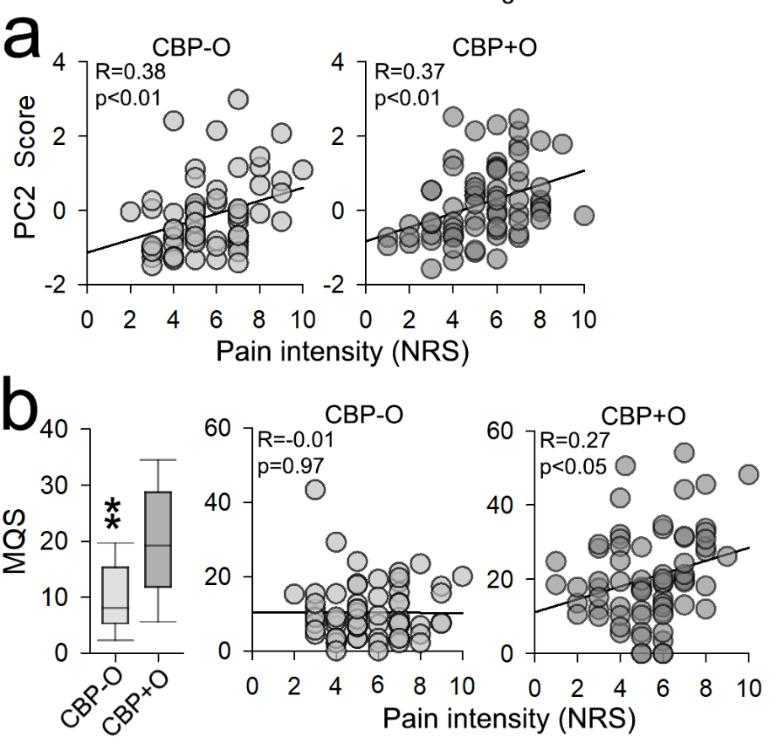

**Extended Data Fig. 2. Relationship of pain intensity with pain quality and with consumption of non-opioid medications, in CBP patients.**

Pain intensity and pain quality (PC2) were similarly positively related in both CBP+O and CBP-O, even though pain quality was worse in CBP+O (a).

CBP+O consumed twice as much non-opioid medications than CBP-O, and in proportion to their pain intensity (b).

These results are informative since even when pain intensity and pain duration are matched between CBP+O and CBP-O, and even though the relationship between pain intensity and pain affect are very similar between the two groups, CBP+O consume more non-opioids in proportion to their pain. The latter provides indirect evidence that long-term opioid use exacerbates the affective component of back pain. Thus, demonstrating an opioid-induced hyper-affective state when pain intensity is matched between the groups, indirectly demonstrating the presence of opioid-induced hyperalgesia.

**a.** PC2 (Pain quality) showed a significant positive relationship with pain intensity for CBP+O and CBP-O patients. **b.** Compared to CBP-O, CBP+O patients showed increased consumption of non-opioid drugs as assessed using MQS (p<0.001). MQS was related to pain intensity in CBP+O group (p<0.01), but not in the CBP-O group (p=0.97). MQS = medication quantification scale. NRS = numerical pain rating scale.

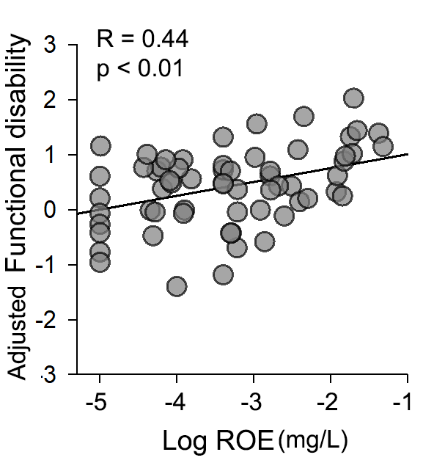

**Extended Data Fig. 3. Opioid blood level is positively associated with worse functional disability (PC1).**

The scatter plot shows the relationship between opioid blood levels (ROE) and PC1 (functional disability) scores corrected for age, sex, NRS, pain duration, BMI, and MQS. BMI = body mass index; MQS = medication quantification scale. NRS = numerical pain rating scale. ROE = relative opioid equivalent in milligrams/liters

The correlation indicates that the higher the blood concentration of opioids, the worse is functional disability in CBP+O.

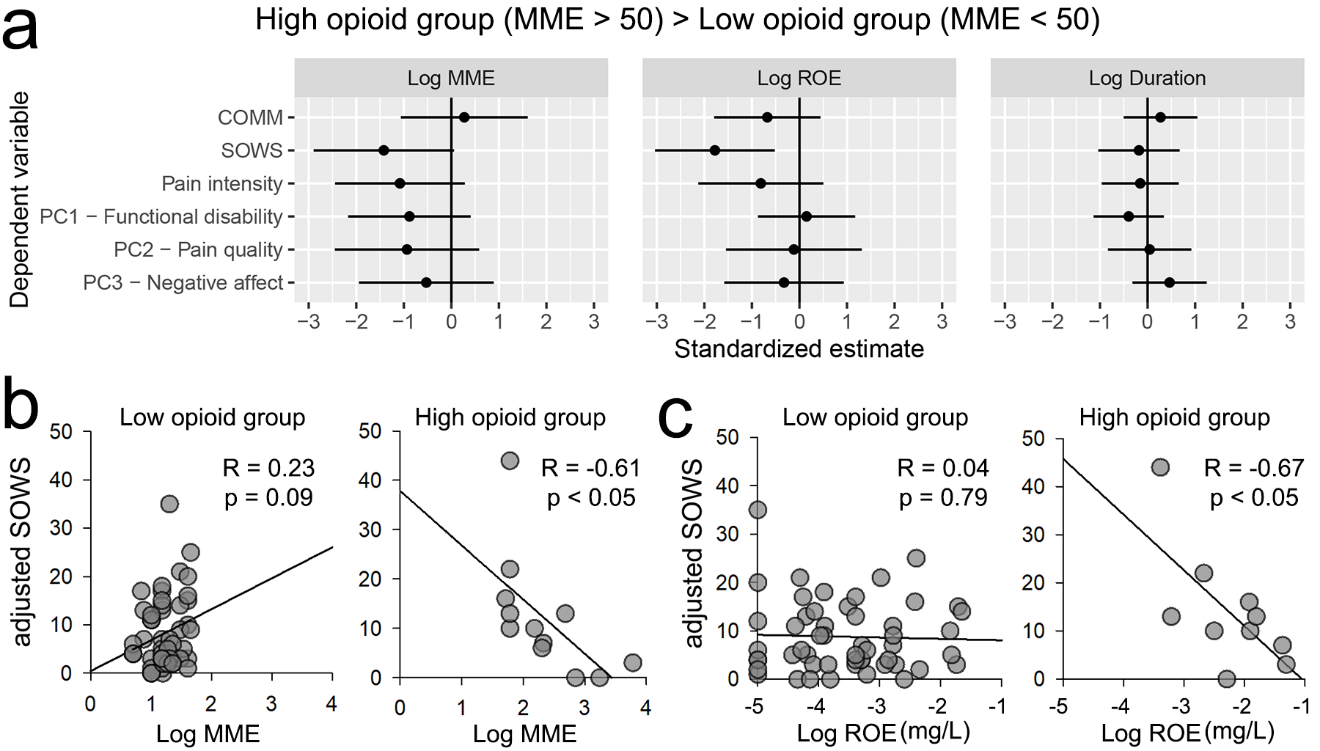

**Extended Data Fig. 4. The relationship between opioid prescription dosage (MME) and signs of withdrawal is dependent on opioid dosage levels when the CBP+O data is subdivided into high (MME > 50) and low opioid consumers (MME < 50).**

Self-assessed opioid withdrawal signs were mostly in the mild to moderate category (mean SOWS score around 10), both in low and high opioid-using subgroups. However, only CBP+O consuming high doses of opioids (MME > 50) show signs of opioid withdrawal (SOWS) dependence on both prescription dosage (MME) and blood levels of opioids (ROE).

**a.** Plot shows standardized estimate differences (± 95% CI) in the relationship between opioid measurements and clinical and psychological outcomes for CBP+O on high opioid dosage (MME ≥ 50, n = 12) and those on low opioid dosage (MME < 50, n =58). MME and ROE showed a more negative relationship with withdrawal symptoms (SOWS) in the high opioid group compared to the low opioid group. **b.** Scatter plots show the relationship between MME and SOWS adjusted for age, sex, pain intensity, pain duration, BMI and MQS in the low opioid group (left scatter) and high opioid group (right scatter). **c**. Scatter plots show the relationship between ROE and SOWS adjusted for age, sex, pain intensity, pain duration, BMI and MQS in the low opioid group (left scatter) and high opioid group (right scatter). BMI = body mass index; MQS = medication quantification scale; NRS = numerical rating scale; MME = daily morphine milligram equivalent; ROE = relative opioid equivalent in milligrams/liters; SOWS = subjective opioid withdrawal scale; COMM = current opioid misuse measurement.

|  | **Peripheral gray matter volume** | | | |
| --- | --- | --- | --- | --- |
|  | **Beta** | **η_p_^2^** | **F-value** | **P-value** |
| ***Opioid (CBP-O > CBP+O)*** | **0.196** | **0.036** | **5.07** | **0.03** |
| ***Sex (Male > Female)*** | **-0.264** | **0.084** | **6.64** | **0.01** |
| ***Race (White > Black)*** | -0.039 | 0.002 | 0.10 | 0.75 |
| ***Age (year)*** | **-0.388** | **0.152** | **19.08** | **<10^-5^** |
| ***BMI*** | -0.098 | 0.012 | 1.25 | 0.27 |
| ***NRS (0-10)*** | 0.079 | 0.008 | 0.63 | 0.43 |
| ***Log Pain duration (year)*** | 0.140 | 0.024 | 2.24 | 0.14 |
| ***MQS*** | -0.057 | 0.003 | 0.32 | 0.57 |
| ***Opioid x Sex*** | 0.022 | 0.001 | 0.05 | 0.82 |
| ***Opioid x Race*** | 0.012 | 0.000 | 0.00 | 0.97 |

**Extended Data Table 5. Analysis of covariance for peripheral gray matter volume with opioid use (CBP-O > CBP+O), demographics (sex, race, age, BMI), and pain (NRS, Log Pain Duration) parameters.**

Long-term opioid use, older age, and females exhibited smaller gray matter volume.

Relative to the effect size we observe for one year of aging, an average of 6 years of opioid consumption decreases gray matter volume by a magnitude equivalent to about 0.5 years of aging.

Effect size and significance of opioid use, demographic and pain properties on peripheral gray matter volume. Significance was determined using ANCOVA. Effect size was computed using partial eta-squared (**η_p_^2^**). **η_p_^2^**<0.05 indicates small effect size; 0.05< **η_p_^2^**B<0.15 indicates medium effect size; **η_p_^2^**> 0.15 indicates large effect size. BMI = body mass index; MQS = medication quantification scale. NRS = numerical rating scale.

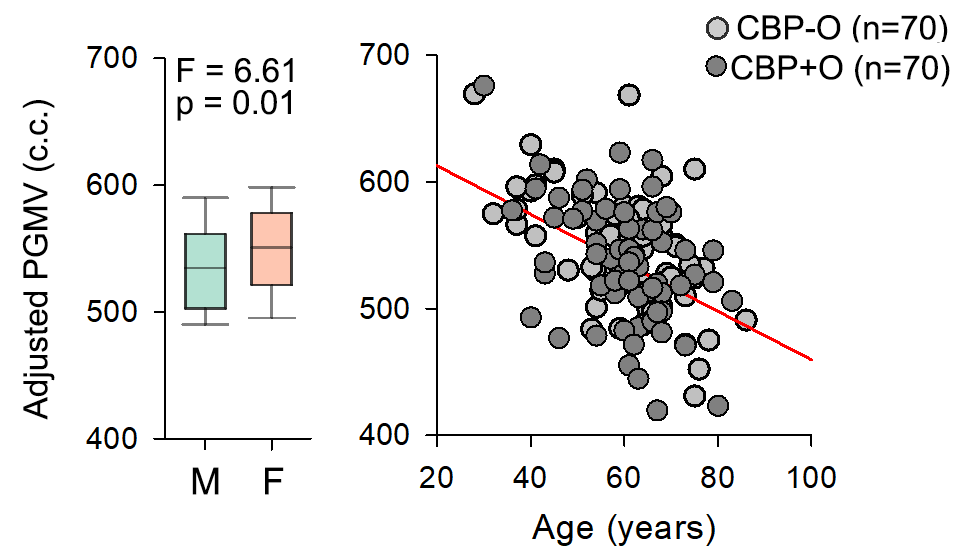

**Extended Data Fig. 5**. **Effect of sex and age on peripheral gray matter volume**

Males showed lower PGMV compared to females (Left box plot). Right scatter plot shows a significant negative relationship between PGMV and age for CBP+O and CBP-O. Statistical values are shown in Extended data Table 5). PGMV = peripheral gray matter volume; M =male; F=female.

|  | **Peripheral gray matter volume** | | | |
| --- | --- | --- | --- | --- |
|  | **Beta** | **η_p_^2^** | **F-value** | **P-value** |
| ***Log MME*** | 0.12 | 0.01 | 0.34 | 0.56 |
| ***Log ROE*** | -0.18 | 0.03 | 0.80 | 0.38 |
| ***Log DOU*** | 0.20 | 0.05 | 1.50 | 0.23 |
| ***SOWS*** | -0.31 | 0.07 | 2.21 | 0.15 |
| ***COMM*** | -0.28 | 0.06 | 2.07 | 0.16 |
| ***PC1- Functional disability*** | 0.02 | 0.00 | 0.02 | 0.88 |
| ***PC2 – Pain quality*** | 0.12 | 0.02 | 0.49 | 0.49 |
| ***PC3 – Negative affect*** | 0.12 | 0.01 | 0.34 | 0.56 |

**Extended Data Table 6. Peripheral gray matter volume in CBP+O (n = 70) does not depend on opioid consumption (MME), blood levels of opioids (ROE), duration of opioid use (DOU), signs of withdrawal (SOWS) or misuse (COMM), and clinical parameters (PC1-3).**

This result suggests that either gray matter volume is an indication of opioid use vulnerability or that it is a consequence of long-term adaptations that have become independent of clinical signs. With the current design, we cannot differentiate between these two options.

Effect size was computed using partial eta-squared (**η_p_^2^**). **η_p_^2^**<0.05 indicates small effect size; 0.05< **η_p_^2^**B<0.15 indicates medium effect size; **η_p_^2^**> 0.15 indicates large effect size indicates small effect size. All regressions were performed after correcting for age, sex, race, BMI, and MQS effects. BMI = body mass index; MQS = medication quantification scale. NRS = numerical rating scale;

|  | **Opioid**  **(CBP-O > CBP+O)** | **Sex**  **(Females > Male)** | **Race**  **(Black > White)** | **Age (years)** | **BMI** | **Pain Intensity** | **Log**  **Pain duration** | **MQS** |
| --- | --- | --- | --- | --- | --- | --- | --- | --- |
| ***Right Thalamus*** | 3.78  -0.20  0.03 | 5.46  -0.21  0.05 | 0.58  -0.07  0.01 | 15.21  -0.37  0.13 | 0.81  -0.08  0.01 | 0.84  -0.08  0.01 | 0.40  0.05  0.01 | 3.43  -0.19  0.03 |
| ***Left Thalamus*** | 1.83  -0.13  0.02 | 5.46  0.20  0.05 | 0.26  0.05  0.00 | 24.19  -0.45  0.19 | 1.34  -0.10  0.01 | 3.25  -0.16  0.03 | 0.12  0.03  0.00 | 1.15  -0.10  0.01 |
| ***Right Caudate*** | 0.00  0.00  0.00 | 7.60  0.27  0.07 | 0.09  0.03  0.00 | 4.48  -0.22  0.04 | 2.04  -0.14  0.02 | 0.00  0.00  0.00 | 0.71  -0.08  0.01 | 0.04  -.02  0.00 |
| ***Left Caudate*** | 1.27  -0.12  0.02 | 4.50  0.18  0.04 | 0.22  0.04  0.00 | 6.71  -0.28  0.23 | 1.79  -0.12  0.01 | 0.81  -0.08  0.01 | 0.10  0.02  0.00 | 0.09  -0.09  0.00 |
| ***Right Putamen*** | 0.99  -0.11  0.01 | 20.79  0.33  0.11 | 0.14  -0.07  0.00 | 16.72  -0.36  0.14 | 0.58  0.04  0.00 | 0.61  -0.07  0.01 | 0.29  0.06  0.00 | 3.05  -0.18  0.03 |
| ***Left Putamen*** | 0.44  -0.06  0.00 | 20.65  0.37  0.18 | 0.37  -0.05  0.00 | 42.47  -0.56  0.30 | 0.03  0.01  0.00 | 3.42  -0.15  0.03 | 0.16  0.03  0.00 | 0.04  0.00  0.00 |
| ***Right Pallidum*** | 1.40  -0.12  0.01 | 1.55  0.11  0.01 | 0.06  -0.02  0.00 | 7.05  -0.26  0.07 | 1.58  0.12  0.02 | 0.07  -0.02  0.00 | 0.06  -0.02  0.00 | 8.78  -0.31  0.08 |
| ***Left Pallidum*** | 0.03  0.00  0.00 | 5.72  0.25  0.06 | 0.18  -0.09  0.01 | 3.06  -0.18  0.03 | 5.35  0.21  0.05 | 0.00  0.00  0.00 | 0.08  -0.02  0.00 | 5.46  -0.21  0.03 |
| ***Right Hippocampus*** | 0.36  -0.07  0.00 | 0.93  -0.09  0.01 | 0.32  -0.06  0.00 | 7.07  -0.27  0.07 | 0.40  -0.06  0.00 | 6.88  -0.25  0.07 | 1.17  -0.11  0.01 | 1.64  -0.14  0.02 |
| ***Left Hippocampus*** | 0.95  -0.08  0.00 | 1.71  -0.13  0.02 | 0.07  0.00  0.00 | 4.40  -0.19  0.04 | 0.00  0.00  0.00 | 3.29  -0.20  0.04 | 0.21  -0.04  0.00 | 0.36  -0.05  0.00 |
| ***Right Amygdala*** | 0.47  0.07  0.00 | 0.01  0.00  0.00 | 6.78  -0.26  0.65 | 0.11  -0.03  0.00 | 0.90  0.09  0.01 | 0.00  0.00  0.00 | 2.99  -0.17  0.03 | 0.16  -0.04  0.00 |
| ***Left Amygdala*** | 1.50  0.13  0.02 | 0.20  -0.04  0.00 | 2.47  -0.16  0.04 | 3.24  -0.17  0.03 | 1.64  0.14  0.02 | 2.57  -0.17  0.04 | 0.00  0.00  0.00 | 1.97  -0.15  0.03 |
| ***Right NAc*** | 3.02  -0.17  0.03 | 0.53  -0.06  0.01 | 0.07  -0.02  0.00 | 35.59  -0.53  0.27 | 0.11  -0.02  0.00 | 3.61  -0.18  0.05 | 0.00  0.00  0.00 | 0.51  -0.07  0.00 |
| ***Left NAc*** | 0.33  -0.05  0.00 | 1.42  0.12  0.02 | 0.08  -0.08  0.01 | 51.88  -0.60  0.31 | 0.02  0.00  0.00 | 1.72  -0.15  0.02 | 0.43  0.06  0.00 | 5.21  -0.20  0.03 |

**Extended Data Table 7. Analysis of covariance for subcortical volumes with opioid use (CBP-O > CBP+O), demographics (sex, race, age, BMI), and pain parameters (intensity, duration, MQS).**

The important outcome is the lack of subcortical volume differences between CBP+O and CBP-O.

The thalamus, caudate, and putamen showed lower volumes in females compared to males.

Right hippocampus volume was negatively associated with pain intensity, while putamen volumes were negatively related to non-opioid medication use.

The right amygdala showed lower volume in White compared to Black.

Data presented as F-value/Beta/effect size (**η_p_^2^**). Shaded cells represent significant effects (p<0.05). Subcortical regions showed no significant differences in volume between CBP+O and CBP-O patients. Most regions exhibited decreased volume in relationship to age. Data presented as. **η_p_^2^**<0.05 indicates small effect size; 0.05< **η_p_^2^**B<0.15 indicates medium effect size; **η_p_^2^**> 0.15 indicates large effect size. BMI = body mass index; MQS = medication quantification scale; NAc = nucleus accumbens.

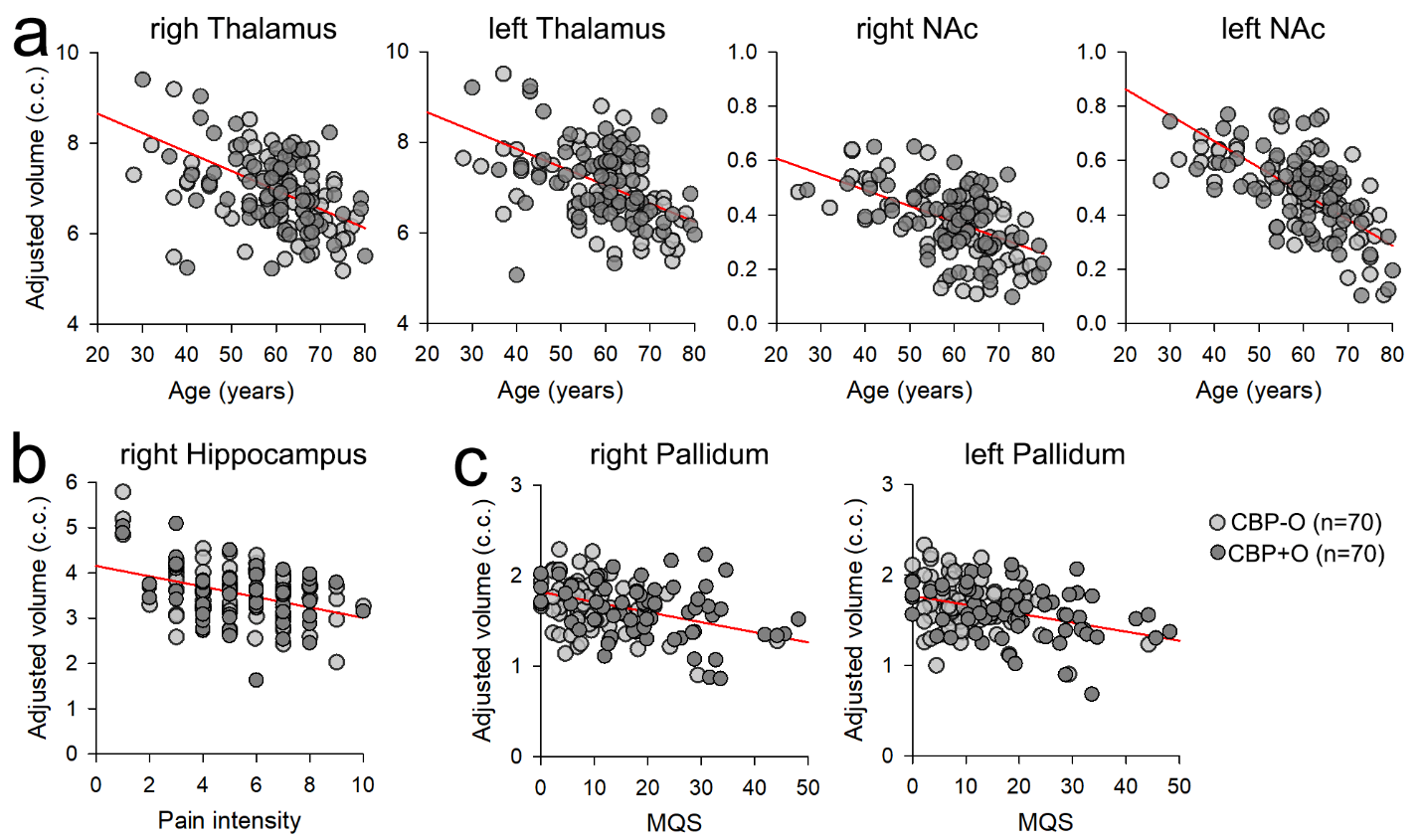

**Extended Data Fig. 6**. **Correlation of subcortical volumes with age, pain, and opioid use.**

**a.** Scatter plots show the top 4 subcortical regions that showed a significant decrease in gray matter volume in relationship to age. Gray matter volumes were adjusted for the other independent variables from the regression model **(see Extended Data Table 7). b.** Right hippocampus volume across all patients was negatively associated with pain intensity. **c.** Bilateral pallidum showed decreased volume with increased non-opioid medication use. Statistics are shown in Extended Data Table 8. NAc = nucleus accumbens.

|  | **Coordinates (mm)** | | | **Size**  **(voxels)** | **T-score** | **Associated terms (Neurosynth reverse inference)** |
| --- | --- | --- | --- | --- | --- | --- |
|  | x | y | z |  |  |  |
| ***CBP-O > CBP+O*** | | | | | | |
| ***mACC*** | 10 | 8 | 38 | 80 | 4.78 | Autonomic control, aversive, pain, noxious, arousal |
| ***Left S1/M1*** | -10 | -32 | 62 | 503 | 5.84 | Sensorimotor, motor function, movement, motor, motor imagery |

**Extended Data Table 8. Regional decreases in gray matter density in patients on long-term opioid use.**

Brain regions that showed significantly decreased gray matter density in CBP+O compared to CPB-O. Terms associated with brain regions were determined using reverse inference from Neurosynth. Coordinates in MNI space; mACC = middle anterior cingulate cortex; S1/M1 = primary sensorimotor cortex.

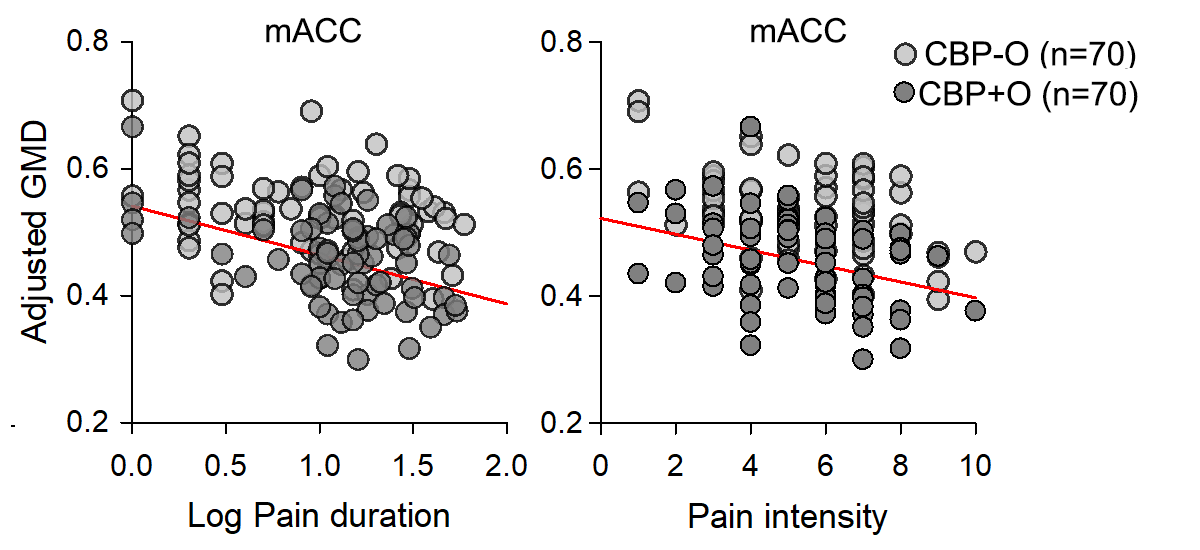

**Extended Data Fig. 7**. **Mid-anterior** **Cingulate Gray matter density was negatively associated with pain log pain duration and pain intensity, in both CBP+O and CBP-O groups.**

Gray matter density in the mACC was significantly associated with pain duration (left scatter plot) and pain intensity (right scatter plot) in all CBP patients (Statistical significance is presented in **Extended data Table 9**). mACC = middle anterior cingulate cortex.

|  | **mACC** | | | | **S1/M1** | | | |
| --- | --- | --- | --- | --- | --- | --- | --- | --- |
|  | **Beta** | **η_p_^2^** | **F-value** | **P-value** | **Beta** | **η_p_^2^** | **F-value** | **P-value** |
| ***Opioid (CBP-O > CBP+O)*** | **0.572** | **0.270** | **36.25** | **<10^-5^** | **0.562** | **0.254** | **33.31** | **<10^-5^** |
| ***Sex (Male > Female)*** | -0.039 | 0.002 | 0.23 | 0.63 | -0.137 | 0.026 | 2.66 | 0.11 |
| ***Race (White > Black)*** | **0.190** | **0.050** | **5.18** | **0.03** | -0.097 | 0.013 | 1.28 | 0.26 |
| ***Age*** | -0.144 | 0.028 | 2.78 | 0.10 | -0.057 | 0.004 | 0.42 | 0.52 |
| ***BMI*** | -0.108 | 0.016 | 1.63 | 0.20 | -0.055 | 0.004 | 0.39 | 0.53 |
| ***NRS*** | **-0.196** | **0.053** | **5.51** | **0.02** | 0.068 | 0.006 | 0.63 | 0.43 |
| ***Log Pain duration*** | **-0.228** | **0.069** | **7.30** | **0.01** | 0.042 | 0.002 | 0.24 | 0.62 |
| ***MQS*** | 0.008 | 0.000 | 0.01 | 0.94 | 0.060 | 0.004 | 0.39 | 0.54 |

**Extended Data Table 9. Analysis of covariance for cortical gray matter density changes (mACC, S1/M1) in patients on long-term opioid use with opioid use (CBP-O > CBP+O), demographics (sex, race, age, BMI), and pain (NRS, Pain duration, MQS) parameters.**

Opioid use, race, pain intensity (NRS), and pain duration were correlated with mACC gray matter density decrease.

Only opioid use was correlated with S1/M1 gray matter density decrease.

Significance was determined using ANCOVA. Effect size was computed using partial eta-squared (**η_p_^2^**). **η_p_^2^**<0.05 indicates small effect size; 0.05< **η_p_^2^**B<0.15 indicates medium effect size; **η_p_^2^**> 0.15 indicates large effect size; BMI = body mass index; MQS = medication quantification scale. NRS = numerical rating scale. mACC = middle anterior cingulate cortex. S1/M1 = primary sensorimotor cortex

|  | **mACC** | | | | **S1/M1** | | | |
| --- | --- | --- | --- | --- | --- | --- | --- | --- |
|  | **Beta** | **η_p_^2^** | **F-value** | **P-value** | **Beta** | **η_p_^2^** | **F-value** | **P-value** |
| ***Log MME*** | 0.13 | 0.01 | 0.39 | 0.54 | 0.04 | 0.00 | 0.04 | 0.85 |
| ***Log ROE*** | -0.23 | 0.04 | 1.23 | 0.28 | 0.14 | 0.01 | 0.40 | 0.53 |
| ***Log DOU*** | 0.06 | 0.00 | 0.15 | 0.70 | -0.05 | 0.00 | 0.08 | 0.78 |
| ***SOWS*** | 0.02 | 0.00 | 0.01 | 0.93 | 0.16 | 0.02 | 0.51 | 0.48 |
| ***COMM*** | 0.05 | 0.00 | 0.07 | 0.79 | -0.07 | 0.00 | 0.12 | 0.73 |
| ***PC1- Functional disability*** | -0.09 | 0.01 | 0.32 | 0.58 | 0.01 | 0.00 | 0.01 | 0.94 |
| ***PC2 – Pain quality*** | 0.12 | 0.02 | 0.49 | 0.49 | -0.19 | 0.03 | 1.05 | 0.31 |
| ***PC3 – Negative affect*** | -0.31 | 0.08 | 2.59 | 0.12 | 0.05 | 0.00 | 0.07 | 0.80 |

**Extended Data Table 10. We observe no relationship between gray matter density for regions decreased in CBP+O (mACC, S1/M1) with opioid use measures (MME, ROE, DOU), clinical parameters (SOWS, COMM), and principal components (PC1-3) in CBP+O patients.**

Effect size was computed using partial eta-squared (**η_p_^2^**). **η_p_^2^**<0.05 indicates small effect size; 0.05< **η_p_^2^**B<0.15 indicates medium effect size; **η_p_^2^**> 0.15 indicates large effect size. All regressions were performed after correcting for age, sex, race, BMI, and MQS effects. BMI = body mass index; MQS = medication quantification scale. NRS = numerical pain rating scale.

|  | **Coordinates (mm)** | | | **Size**  **(voxels)** | **T-score** | **Associated terms (Neurosynth reverse inference)** |
| --- | --- | --- | --- | --- | --- | --- |
|  | x | y | z |  |  |  |
| ***Regions that exhibited increased ALFF in CBP+O (CBP+O > CBP-O)*** | | | | | | |
| ***right pITG*** | 56 | -50 | -8 | 325 | 4.01 | Expression, facial, response time, perception, threating |
| ***left pITG*** | -54 | -50 | -14 | 410 | 4.22 | Words, form, visual words, subsequent memory, orthographic |
| ***mPFC*** | 4 | 60 | 2 | 823 | 6.01 | Autobiographical, default-mode, cognitive, intention |
| ***right LOC*** | 26 | -64 | 52 | 910 | 5.45 | Attention, spatial, calculation, rotation, orientation |
| ***MCL*** | 2 | 20 | -10 | 5050 | 7.89 | Reward, value, motivation, monetary, incentive |
| *right NAC* | 8 | 10 | -8 | 164 | 7.13 |  |
| *left NAc* | -10 | 10 | -10 | 162 | 7.05 |  |
| *sgACC* | 2 | 24 | -14 | 516 | 6.85 |  |
| *aACC* | -2 | 34 | -4 | 404 | 6.11 |  |
| *right Amyg* | 18 | -2 | -18 | 44 | 4.01 |  |
| *left Amyg* | -20 | -2 | -20 | 28 | 3.79 |  |
| *Brainstem* | -2 | -26 | -8 | 86 | 3.25 |  |
| ***Regions that exhibited decreased ALFF in CBP+O (CBP+O < CBP-O)*** | | | | | | |
| ***right aMTG*** | 54 | -8 | -20 | 1275 | -6.58 | Mind, mental states, theory, social, experiences |
| ***left aMTG*** | -54 | -8 | -22 | 1309 | -6.94 | Semantic, sentences, mentalizing, linguistic, theory of mind |
| ***left dlPFC*** | -46 | 24 | 22 | 3955 | -5.08 | Language, phonological, syntactic, verb, demands |
| *left IFG* | -50 | 15 | 6 | 1478 | -5.06 |  |
| *left MFG* | -42 | 30 | 32 | 2040 | -5.02 |  |

**Extended Data Table 11. Regional ALFF changes in patients on long-term opioid use.**

The table shows Coordinates in mm (MNI space), cluster size and T-score of brain regions that showed significant differences in ALFF between CBP+O and CBP-O (Threshold free Cluster corrected, T-score > 2.3, p < 0.01). The local coordinates, sizes, and T-scores for distinct subregions within the MCL and dlPFC clusters are shown. The top 5 associated terms with any given cluster were determined using reverse inference from Neurosynth.

The ALFF = Amplitude of low frequency fluctuations; pITG = posterior inferior temporal gyrus; MCL = meso-cortical limbic. mPFC = middle prefrontal cortex; LOC = Lateral occipital cortex; aMTG = anterior middle temporal gyrus; dlPFC = dorsolateral prefrontal cortex. Amyg = amygdala; NAc = Nucleus accumbens; sgACC = subgenual division of anterior cingulate cortex; aACC = anterior division of division of anterior cingulate cortex; IFG = Inferior frontal gyrus; MFG = Middle frontal gyrus.

|  | **Log MME**  R-value  (P-value) | **Log ROE**  R-value  (P-value) | **Log DOU**  R-value  (P-value) | **SOWS**  R-value  (P-value) | **COMM**  R-value  (P-value) | **PC1**  R-value  (P-value) | **PC2**  R-value  (P-value) | **PC3**  R-value  (P-value) |
| --- | --- | --- | --- | --- | --- | --- | --- | --- |
| ***Regions that exhibited increased ALFF (CBP+O > CBP-O)*** | | | | | | | | |
| ***right pITG*** | 0.13 (0.33) | -0.05 (0.69) | -0.17 (0.19) | 0.08 (0.52) | -0.04 (0.76) | 0.10 (0.40) | -0.02 (0.85) | 0.12 (0.37) |
| ***left pITG*** | 0.04 (0.76) | 0.03 (0.79) | -0.15 (0.26) | -0.03 (0.79) | 0.11 (0.40) | 0.20 (0.10) | 0.06 (0.64) | 0.11 (0.39) |
| ***mPFC*** | -0.05 (0.67) | -0.07 (0.60) | -0.09 (0.50) | 0.04 (0.76) | 0.08 (0.52) | 0.08 (0.52) | 0.08 (0.53) | 0.15 (0.26) |
| ***right LOC*** | 0.21 (0.09) | 0.22 (0.08) | -0.05 (0.68) | 0.11 (0.39) | 0.13 (0.35) | 0.10 (0.41) | 0.10 (0.40) | 0.01 (0.93) |
| ***MCL*** | 0.12 (0.38) | 0.01 (0.97) | -0.04 (0.74) | 0.10 (0.40) | -0.04 (0.76) | 0.16 (0.27) | 0.05 (0.68) | 0.14 (0.29) |
| ***Regions that exhibited decreased ALFF (CBP+O < CBP-O)*** | | | | | | | | |
| ***right aMTG*** | -0.15(0.28) | -0.03 (0.81) | 0.02 (0.89) | 0.14 (0.30) | -0.02 (0.86) | -0.04 (0.76) | 0.01 (0.94) | 0.02 (0.86) |
| ***left aMTG*** | -0.02 (0.86) | 0.08 (0.52) | -0.01 (0.93) | 0.07 (0.59) | 0.07 (0.59) | -0.07 (0.59) | 0.06 (0.69) | 0.06 (0.69) |
| ***left dlPFC*** | -0.22 (0.08) | -0.02 (0.88) | -0.01 (0.93) | -0.04 (0.74) | 0.02 (0.87) | -0.13 (0.33) | -0.19 (0.10) | 0.04 (0.76) |

**Extended Data Table 12. Brain regional activity (ALFF) changes between CBP+O and CBP-O were not correlated with opioid use (MME, ROE, DOU), clinical parameters (SOWS, COMM), and characteristics of back pain (PC1-3), in CBP+O patients.**

Data show the correlation of ALFF with MME, ROE, DOU, COMM, SOWS, PC1-functional disability, PC2-pain quality, and PC3-negative affect of for regions that showed significant ALFF differences between CBP+O and CBP-O. ALFF values were corrected for age, sex, race, pain intensity, BMI and MQS prior to correlation. ALFF = Amplitude of low-frequency fluctuations; pITG = posterior inferior temporal gyrus; MCL = meso-cortical limbic; mPFC = middle prefrontal cortex; LOC = Lateral occipital cortex; aMTG = anterior middle temporal gyrus; dlPFC = dorsolateral prefrontal cortex. MME = Morphine Milligram Equivalents; ROE = Relative Opioid Equivalent; DOU = duration of opioid use; SOWS = subjective opioid withdrawal scale; COMM = current opioid misuse measure.

|  | **CBP+O (MME>=50)**  (mean ± s.d.) | **CBP+O (MME<50)**  (mean ± s.d.) | **T-value** | **p-value** |
| --- | --- | --- | --- | --- |
| ***Regions that exhibited increased ALFF (CBP+O > CBP-O)*** | | | | |
| ***right pITG*** | 1.04 ± 0.02 | 1.03 ± 0.02 | 0.32 | 0.752 |
| ***left pITG*** | 1.07 ± 0.03 | 1.07 ± 0.03 | -0.17 | 0.867 |
| ***mPFC*** | 0.98 ± 0.02 | 1.00± 0.02 | -1.46 | 0.146 |
| ***right LOC*** | 1.04 ± 0.03 | 1.03 ± 0.02 | 1.29 | 0.199 |
| ***MCL*** | 1.03 ± 0.02 | 1.04 ± 0.03 | -1.02 | 0.318 |
| ***Regions that exhibited decreased ALFF (CBP+O < CBP-O)*** | | | | |
| ***right aMTG*** | 0.99 ± 0.02 | 0.98± 0.02 | 0.05 | 0.996 |
| ***left aMTG*** | 0.98 ± 0.02 | 0.97± 0.01 | 1.33 | 0.187 |
| ***left dlPFC*** | 0.97 ± 0.02 | 0.96 ± 0.02 | 0.763 | 0.448 |

**Extended Data Table 13. Regional activity (ALFF) changes in CBP+O did not differ between low (MME<50) and high (MME>50) opioid consumption.**

Data indicate differences in ALFF ROI between CBP+O patients with high MME (MME >= 50) and those with low MME (MME < 50). There were no differences between groups for all regions examined. ALFF = Amplitude of low-frequency fluctuations; pITG = posterior inferior temporal gyrus; mPFC = middle prefrontal cortex; LOC = Lateral occipital cortex; aMTG = anterior middle temporal gyrus; dlPFC = dorsolateral prefrontal cortex. MME = Morphine Milligram Equivalents; ROE = Relative Opioid Equivalents; BMI = body mass index; MQS = medication quantification scale.

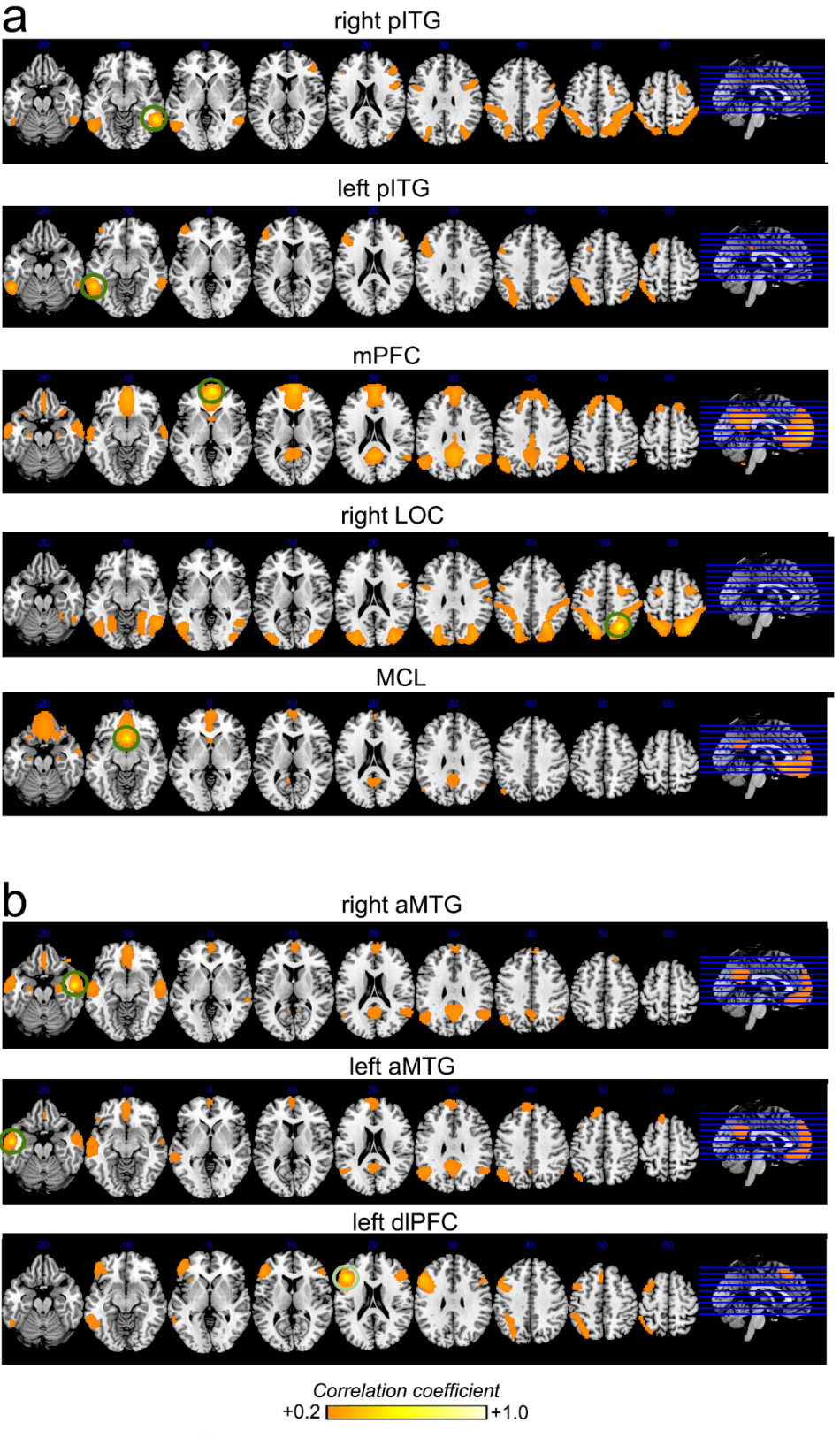

**Extended Data Fig. 8**. **Resting-state functional connectivity maps identified for brain regions that showed activity (ALFF) changes with long-term opioid use.**

Each of the eight regions, identified from the ALFF contrast between CBP+O and CBP-O, when used as a seed in 846 healthy subjects resting state fMRI (Connectome 1000) mapped into a specific canonical resting state network: 1. Right pITG seed identifies the right executive control network; 2. Left pITG identifies the left executive control network; 3. The mPFC seed identifies the default mode network (DMN); 4. The LOC seed identifies both dorsal attention and executive control networks; 5. The MCL seed identifies limbic structures and parts of DMN; 6. Left and right MTG identify the DMN; 7. Left dlPFC identifies the left executive function network.

Brain slices represent the group mean resting state functional correlation maps from 846 healthy subjects for regions that showed increased **(a)** and decreased **(b)** ALFF in CBP+O compared to CBP-O. Maps were generated using seed-based region of interest analysis. Regions used to generate each map are delineated by green circles. Connectivity maps are displayed thresholded at R-value > 0.2 for clarity. ALFF = Amplitude of low-frequency fluctuations; pITG = posterior inferior temporal gyrus; mPFC = middle prefrontal cortex; LOC = Lateral occipital cortex; aMTG = anterior, middle temporal gyrus; dlPFC = dorsolateral prefrontal cortex; MCL = mesocorticolimbic region.

| **Regressor** | **CBP-O (n=70)**  DMN+MCL dwell times (sec)  mean ± s.d. | **CBP+O (n=70)**  DMN+MCL dwell times (sec)  mean ± s.d. | **T-score (P-value)** |
| --- | --- | --- | --- |
| ***None*** | 7.78 ± 1.16 | 8.57 ± 1.48 | -3.53 (<10^-2^) |
| ***right pITG*** | 7.59 ± 1.16 | 8.04 ± 1.34 | -2.13 (0.03) |
| ***left pITG*** | 7.61 ± 1.17 | 8.11 ± 1.39 | -2.27 (0.02) |
| ***mPFC*** | 7.10 ± 0.97 | 7.52 ± 1.31 | -2.27 (0.02) |
| ***right LOC*** | 7.46 ± 1.02 | 7.99 ± 1.39 | -2.04 (0.05) |
| ***MCL*** | 7.61± 1.12 | 7.87 ± 1.31 | -1.24 (0.21) |
| ***right aMTG*** | 7.43 ± 1.01 | 7.85 ± 1.27 | -2.11 (0.03) |
| ***left aMTG*** | 7.13± 0.99 | 7.72 ± 1.23 | -3.15 (<10^-2^) |
| ***left dlPFC*** | 7.75 ± 1.22 | 8.19 ± 1.35 | -2.04 (0.05) |

**Extended Data Table 14. Long-term opioid use was associated with increased dwell times in the DMN+MCL network, primarily mediated by MCL activity.**

CBP+O showed significantly higher dwell times in the DMN+MCL network (first row).

This difference disappears when regressing out BOLD signal from MCL region but is maintained when regressing out the BOLD signal from the other seven regions. Therefore, the MCL seems the main controller of dwell time for the DMN+MCL network.

Differences in means were determined using an unpaired t-test after correcting the dwell times for age, sex, BMI, MQS, NRS and pain intensity. pITG = posterior inferior temporal gyrus; mPFC = middle prefrontal cortex; LOC = Lateral occipital cortex; MCL = meso-cortical limbic; aMTG = anterior middle temporal gyrus; dlPFC = dorsolateral prefrontal cortex; DMN = Default-mode network; MCL = Mesocorticolimbic

|  | **CBP-O (n=46)**  mean ± s.d. | **CBP+O (n=22)**  mean ± s.d. | **p-value** |
| --- | --- | --- | --- |
| ***Demographic characteristics*** | | | |
| Age (years) | 64.09 ± 8.72 | 59.72 ± 10.68 | 0.12 |
| Sex/female (%) | 30 (65.31%) | 15 (68.18%) | 0.84 |
| ***Pain characteristics*** | | | |
| Pain intensity (NRS) | 6.36 ± 1.96 | 6.63 ± 1.97 | 0.62 |
| Pain duration (years) | 14.54 ± 17.22 | 15.91± 14.44 | 0.68 |

**Extended data Table 15. Demographics and pain characteristics for a subgroup of 44 CBP-O and 22 CBP+O patients where NIH toolbox measurements and resting state fMRI data were available.**

CBP+O and CBP-O subgroups were group-matched for age, sex, pain intensity, and duration.

Data presented as mean ± s.d. and was obtained from our previous study (https://doi.org/10.1101/2022.09.14.22279907).

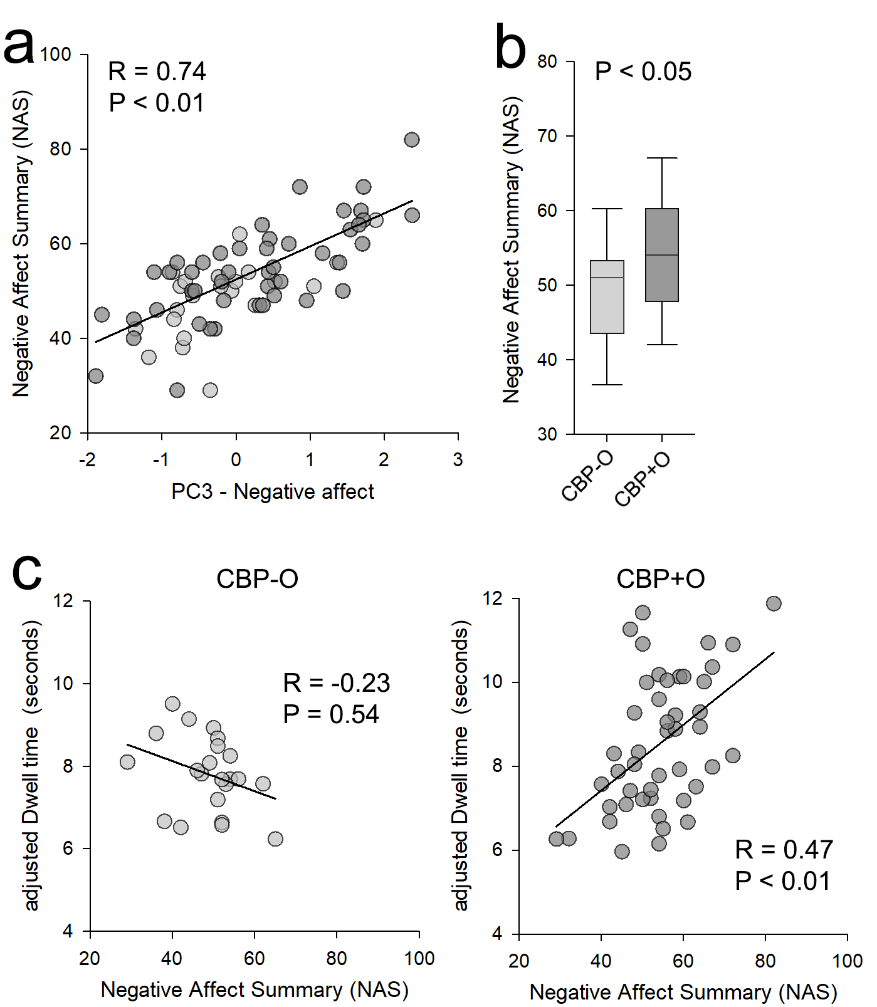

**Extended Data Fig. 9**. **The relationship of dwell times of the DMN+MCL network with negative affect was consistent across two negative affect measurements.**

**a.** Negative Affect Summary (NAS) score (from NIH Toolbox) showed a strong positive association with PC3-Negative affect for a subgroup of CBP+O (n =46) and CBP-O (n=22) patients. **b.** CBP+O patients showed higher NAS scores compared to CBP-O. **c.** NAS was positively associated with dwell times of the DMN+MCL network in CBP+O but not in CBP-O patients.

|  | **Baseline**  (mean ± s.e.m.) | **Abstinence**  (mean ± s.e.m) | **T-value** | **p-value** |
| --- | --- | --- | --- | --- |
| ***Log ROE (mg/L)*** | -3.42 ± 0.51 | -4.51 ± 0.29 | 4.57 | <0.001 |
| ***Pain intensity (NRS)*** | 5.07 ± 0.51 | 5.28 ± 0.79 | -0.42 | 0.68 |
| ***SF12 physical*** | 29.62 ± 1.30 | 30.31 ± 1.35 | -1.79 | 0.09 |
| ***SF12 mental*** | 51.88 ± 3.34 | 46.97 ± 2.58 | 3.99 | <0.01 |
| ***SOWS*** | 6.14 ± 1.22 | 7.57 ± 0.03 | -2.16 | <0.05 |

**Extended Data Table 16. Behavioral changes between baseline and following a brief period of opioid abstinence in 14 patients on long-term opioids.**

Blood levels of opioids (Log ROE) were significantly lower after opioid abstinence. Opioid abstinence results in decreased SF12 mental function scores and increased withdrawal signs. No changes were observed for pain intensity or SF12 physical function (paired t-test). NRS = Numerical Rating Scale; ROE = Relative Opioid Equivalents; SOWS = subjective opioid withdrawal scale.

|  | **Baseline**  (mean ± s.d.) | **Abstinence**  (mean ± s.d.) | **T-value** | **p-value** |
| --- | --- | --- | --- | --- |
| ***Regions that exhibited increased ALFF (CBP+O > CBP-O)*** | | | | |
| ***right pITG*** | 1.04 ± 0.02 | 1.06 ± 0.02 | -1.92 | 0.08 |
| ***left pITG*** | **1.07 ± 0.03** | **1.03 ± 0.03** | **3.39** | **0.004*** |
| ***mPFC*** | 1.01 ± 0.02 | 0.96 ± 0.02 | 1.89 | 0.09 |
| ***right LOC*** | 1.03 ± 0.03 | 1.04 ± 0.03 | -1.28 | 0.15 |
| ***MCL*** | 1.04 ± 0.04 | 1.02 ± 0.03 | 1.22 | 0.24 |
| ***Regions that exhibited decreased ALFF (CBP+O < CBP-O)*** | | | | |
| ***right aMTG*** | **0.99 ± 0.03** | **0.89± 0.04** | **6.78** | **<10^-4^ *** |
| ***left aMTG*** | 0.96 ± 0.03 | 0.94 ± 0.02 | 1.84 | 0.09 |
| ***left dlPFC*** | 0.97 ± 0.02 | 0.98 ± 0.01 | -0.95 | 0.36 |

**Extended Data Table 17. Regional ALFF changes, between before and following opioid abstinence, in 14 patients on long-term opioids.**

Most brain regions that showed significant ALFF differences between CBP+O and CBP-O showed minimal changes following opioid abstinence. Only the left pITG and right aMTG showed any significant change (paired t-test p<0.05). ALFF = Amplitude of low-frequency fluctuations; pITG = posterior inferior temporal gyrus; mPFC = middle prefrontal cortex; LOC = Lateral occipital cortex; MCL = mesocorticolimbic region; aMTG = anterior middle temporal gyrus; dlPFC = dorsolateral prefrontal cortex. *p<0.05 Bonferroni corrected.
